# Dengue clinical features and predictors of severity in the diabetic patient: a retrospective cohort study on Reunion island, 2019

**DOI:** 10.1101/2023.02.27.23286123

**Authors:** Azizah Issop, Antoine Bertolotti, Yves-Marie Diarra, Jean-christophe Maïza, Éric Jarlet, Muriel Cogne, Éric Doussiet, Éric Magny, Olivier Maillard, Epidengue Cohort Investigation Team, Estelle Nobécourt, Patrick Gérardin

## Abstract

Aim: Diabetes mellitus is associated with both the risks of severe dengue and dengue-related deaths, however the factors characterizing dengue in the diabetic patient are ill-recognized. The objective of this hospital-based cohort study was to identify the factors characterizing dengue and those able to predict dengue severity in the diabetic patient.

**Methods:** We retrospectively analysed demographic, clinical and biological parameters at admission in the cohort of patients who consulted at the university hospital between January and June 2019 with confirmed dengue. Bivariate and multivariate analyses were conducted.

**Results:** Of 936 patients, 184 patients (20%) were diabetic. One hundred and eighty-eight patients (20%) developed severe dengue according to the WHO 2009 definition. Diabetic patients were older and had more comorbidities than non-diabetics. In an age-adjusted logistic regression model, loss of appetite, altered mental status, high neutrophil to platelet ratios (>14.7), low haematocrit (≤ 38%), upper-range serum creatinine (>100 µmol/l) and high urea to creatinine ratio (>50) were indicative of dengue in the diabetic patient. In a modified Poisson regression model, four key independent variables were predictive of severe dengue in the diabetic patient: presence of diabetes complications, non-severe bleeding, altered mental status and cough. Among diabetes complications, diabetic retinopathy and neuropathy, but not diabetic nephropathy nor diabetic foot, were predictive of severe dengue.

**Conclusion:** At hospital first presentation, dengue in the diabetic patient is characterized by deteriorations in appetite, mental and renal functioning, while severe dengue can be predicted by presence of diabetes complications, dengue-related non-severe haemorrhages, cough, and dengue-related encephalopathy.

## Introduction

Dengue virus (DENV) is the most common arbovirus circulating in the world [1]. In 2022, the European Center for Disease Prevention and Control (ECDC), reported from different sources 4,110,465 dengue cases and 4,099 dengue-related deaths worldwide. DENV affects mainly tropical countries of South America, Asia, Africa and Western Pacific [2]. However, with the spread of the *Aedes albopictus* vector and the sharp increase in traveller flows, it is no longer limited to the inter-tropical zone and dengue epidemics are also documented in temperate regions [3]. In metropolitan France, where *Aedes albopictus* is established and spreads since 2004, episodes of autochthonous dengue transmission occur since 2010[4, 5].

Reunion island, a French overseas department located in the south-western Indian ocean (SWIO) region, is particularly affected by DENV, which has been circulating under an epidemic mode since 2018. Thus, despite *Ae albopictus* related transmission may be considered as a distinct outlier associated with slow, low intensity outbreaks [6], 18,217 autochthonous cases have been notified to the island surveillance system in 2019, of which 1,944 occasioned visits to emergency wards and 620 ended in hospitalization [7].

The infection results in a wide spectrum of clinical manifestations that range from mild asymptomatic or paucisymptomatic infections, undifferentiated fever, dengue fever (DF) to severe dengue (SD), which is currently designated as severe plasma leakage, severe bleeding and/or severe organ involvement [8].

Predicting the occurrence of severe dengue in patients at risk remains a major public health challenge. Overall, diabetes mellitus is known to increase the susceptibility to infection [9] and diabetic patients tend to have a higher risk for infection-related mortality through major cardiovascular events such as myocardial infarction and stroke [10, 11]. In addition, diabetes was reported as a factor that drives the progression from DF to SD [12, 13]. With respect to dengue, diabetes is therefore of paramount importance, and all the more so in Reunion island, which is one of the French region with the highest prevalence of treated diabetes (10% of the adult population, *i.e.*, twice the national average [14]).

The primary objective of this retrospective cohort study was twofold and aimed to identify both the factors characterizing dengue and those able to predict severe dengue in the diabetic patient among the subjects presenting at the hospital. The secondary objectives were to distinguish the clinical features of dengue among the subjects presenting with uncomplicated or complicated diabetes or presenting with optimal or suboptimal glycaemic control (glycated haemoglobin, HbA1c <7% or ≥ 7%).

## Patients and methods

### Setting and population

We retrospectively enrolled in the cohort all patients with suspected dengue who consulted at the university hospital between January 1 and June 30, 2019, and decided to restrict the analyses to virologically confirmed cases of dengue.

Subjects were suspected of being infected with DENV if they had fever higher than 38°C and at least one of the following symptoms: headache, retro-orbital pain, myalgia, arthralgia, rash, or pruritus. Dengue was confirmed if the patient had a positive detection of DENV RNA by reverse transcriptase chain reaction (rt-PCR) and/or a positive detection of non-specific antigen 1 (NS1) by Rapid Diagnostic Test (RDT). The diagnosis of SD was made when the World Health Organization (WHO) 2009 criteria were met. Briefly, SD was diagnosed in presence of: (1) severe plasma leakage leading to shock or respiratory distress, (2) severe bleeding, or (3) severe organ involvement (*e.g.*, high liver enzyme levels AST ALT ≥1000 IU/L, impaired consciousness, heart and other severe organ dysfunctions) [8]. The criteria used for defining SD and the criteria used for defining severe organ involvement in SD are detailed in the methodological appendix (S0 Table).

We systematically reviewed the medical records of all patients over 15 years admitted to the University Hospital of Reunion island for a clinical suspicion of dengue infection. We analysed in the cohort only the subjects with a confirmed diagnosis of dengue. For each patient with a confirmed dengue, it was verified that the hospital visit was related to the DENV infection. Thus, the patients hospitalized for another cause were ruled out.

We retrieved socio-demographic, clinical and biological data for each patient in the EPIDENGUE database, an electronic record based on the cross-referencing of data from the PMSI (*Programme de Médicalisation des Système d’Information*) and the laboratories completed by junior doctors. The biological data were collected at hospital first consultation.

Diabetes history was reported if documented and registered in the medical file and/or the patient received an anti-diabetic treatment.

### Statistical analysis

For bivariate analysis, quantitative variables were expressed as medians and interquartile ranges (1^st^ - 3^rd^ quartiles). Qualitative variables were expressed as numbers and percentages. Percentages were compared between diabetic patients (DPs) and non-diabetic patients (NDPs), and between severe and non-severe dengue cases using Chi-square and Fisher exact tests, as appropriate. Medians were compared between these groups using non-parametric Brown-Mood tests [15]. Medians that were statistically different were further tested using median and quartile regressions to see whether they differ substantially both in precision and dispersion [16].

Subgroup analyses were conducted under the same modalities to assess both the influence of complicated diabetes and of the control of blood sugar on dengue presentation.

Multivariate analyses were conducted first to identify the clinical and biological variables associated with diabetes among patients with dengue, second to find predictors of SD in the DP. In the first model, we used a non-conditional logistic regression model for estimating odds ratios (OR) and 95% confidence intervals (95%CI) of the factors independently associated with diabetes among the patients attending the hospital with dengue.

In the second model, we used the adapted Poisson regression model for binary outcome with robust variance option [17] for estimating the relative risks (RR) and 95%CI of the predictors of severe dengue in the DPs. In both analyses, we set *a priori* P, the maximum number of candidate variables (parameters) to be selected using the formula:

P = exp <u>(0.508 – 0.259 ln (</u>□<u>) + 0,544 ln(</u>□<u>) + ln(MAPE)/0.504),</u> wherein □ = is the proportion of the outcome, n is the population size and MAPE, the mean absolute prediction error [18]. In agreement with this method, we began with full models controlling all variables with *p* values <0.025 in bivariate analyses (with socio-demographic/medical history and clinical data separate; clinical and biological data separate, respectively). Each model was added to the other in an intermediate joined model (clinico-biological, or clinico-anamnestic, respectively) which was submitted to a stepwise backward procedure that aimed at eliminating the less significant variables. Final parsimonious models retained only significant covariates. However, as the prevalence of diabetes increases with age, we forced age in the first final model. The fitness of the different models was evaluated using the Bayesian Information Criterion (BIC). All interaction terms between the covariates included in the final models were tested, likewise the effects of age and sex as potential disease modifiers. Model calibration was evaluated using the Hosmer-Lemeshow goodness-of-fit test (also applied to the corresponding logistic model, when not applicable for Poisson regression models). The discriminative capacity of the models was evaluated by the area under the receiver operating characteristic curve (AUC). Analyses were performed using Stata® (v16.1, StataCorp, College Station, Tx, 2019). For all these, observations with missing data were ruled out and a two-tailed *p* value <0.05 was considered significant.

### Ethical approval

This monocentric observational retrospective study was conducted according to the reference methodology MR-004 from *the National Commission of Informatics and Liberties*. In accordance with French regulations, this retrospective study did not require approval from an ethics committee. The ethical character of the study on previously collected data was approved by the Institutional Review Board of the *Centre Hospitalier Universitaire Reunion*. The EPIDENGUE database was registered in the national health data hub (n° F20201021104344). Patients were informed of the study and non-refusal of participation was collected. Data was treated anonymously from patient’s medical records.

## Results

### Dengue in the diabetic and non-diabetic patients

Between January 1, 2019, and the June 30, 2019, the surveillance system reported more than 42,000 dengue illnesses on Reunion island, of which up to 18,000 were biologically confirmed (>30,000 and >13,000 for the southern region alone, respectively). During this period, the EPIDENGUE database enrolled 2,365 subjects clinically suspected of dengue having presented at the two centres of the teaching hospital among whom this study included 936 patients with confirmed DENV infection (Fig. 1). Of these 936 patients, 184 were diabetic and 752 non-diabetic. We did not distinguish between type 1 and 2 diabetes, given the small number of patients primarily treated with insulin alone.

**Figure 1.**
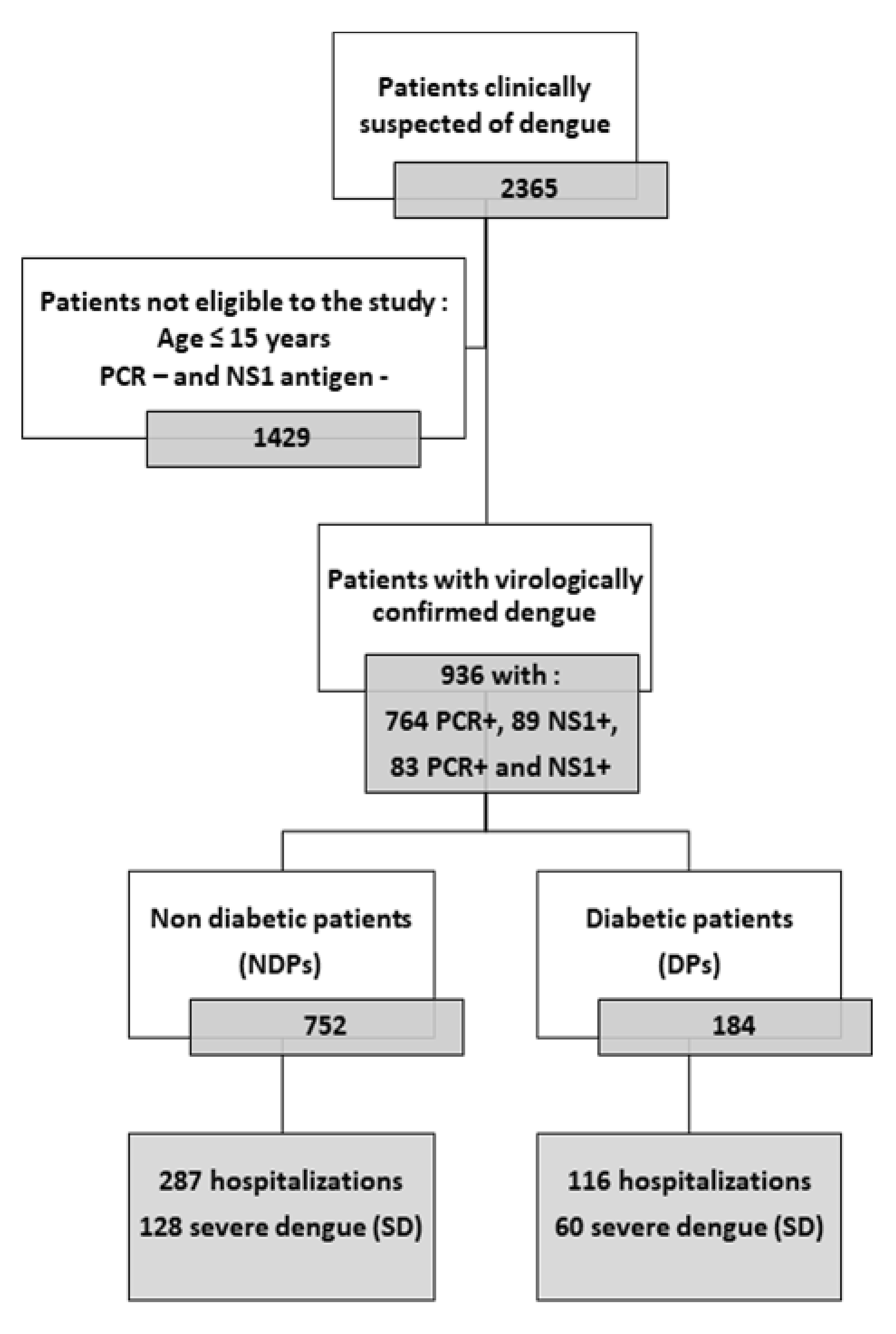
Study population

The most important differences between DPs and NDPs are presented in Table 1, while the full comparison between both populations is displayed in S1 Table.

**Table 1.**
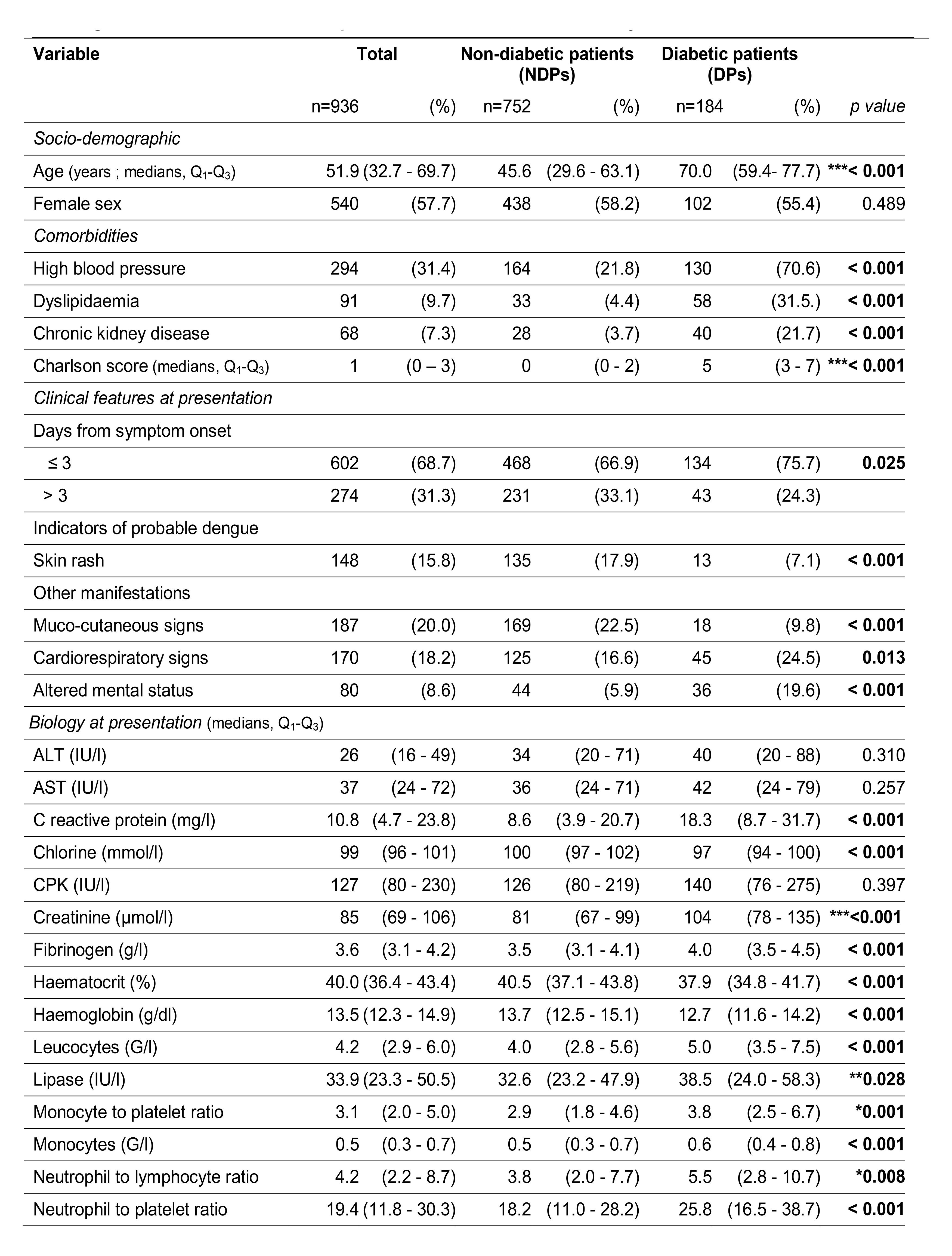
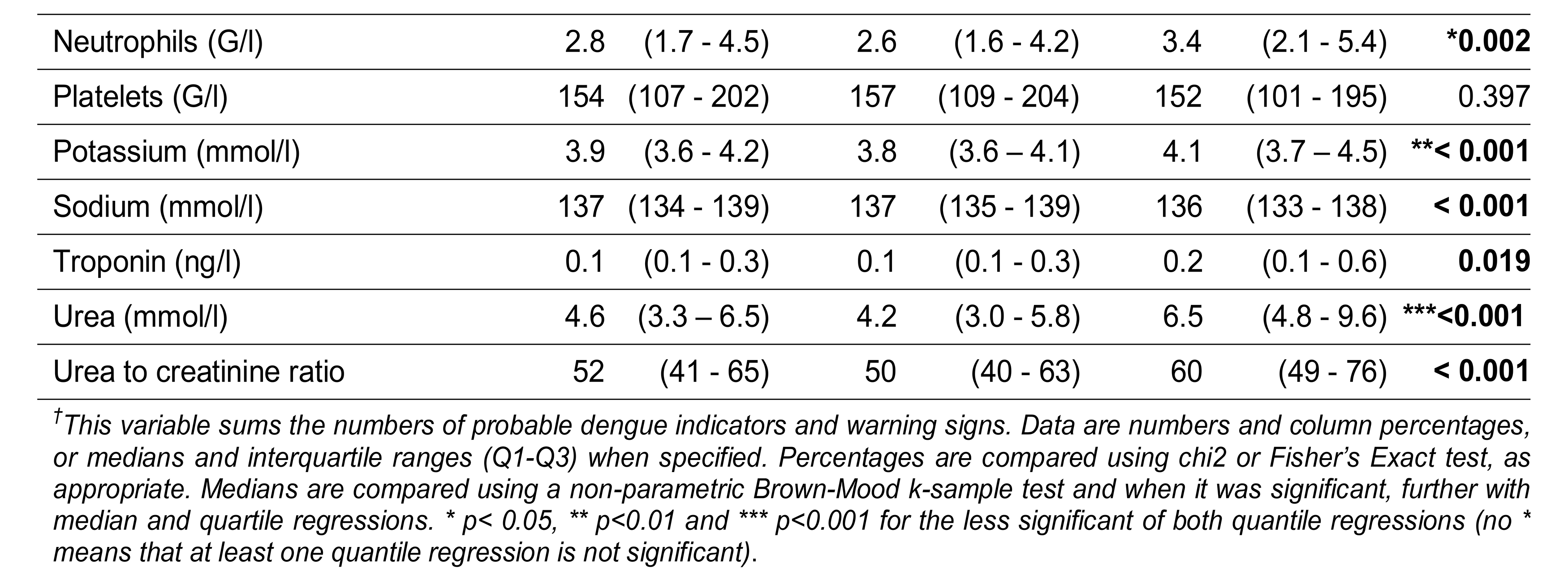
Characteristics at hospital presentation of 936 virologically confirmed cases of dengue among non-diabetic and diabetic patients, Reunion island, January to June 2019.

DPs were older than NDPs (median age 70.0 years; Q_1_-Q_3_: 59.4-77.7 *versus* 45.6 years, Q_1_-Q_3_: 29.7-63.0, *p*<0.001). The proportion of women was similar in both populations. Comorbidities were more frequent in DPs and ranged between 5.4% for peptic ulcer and 70.6% for hypertension (p<0.01 for all comparisons), which resulted in higher Charlson comorbidity index scores among DPs than among NDPs (median score 5; Q_1_-Q_3_: 3-7 *vs* 0, Q_1_- Q_3_: 0-3, *p*<0.001) and more common use of related medications (S1 Table).

DPs reported more frequently fever, fatigue, loss of appetite, and less frequently myalgia within the days preceding first hospital presentation than NDPs. At first consultation, they were more febrile, more tachycardic and exhibited higher systolic and mean blood pressures than NDPs but the differences were not substantial to make sense clinically, as evidenced by inconsistent quantile regression coefficients.

Over the course of the dengue episode, DPs presented earlier to hospital than NDPs (75.7% *vs* 66.9%, within 3 days of symptom onset, p=0.025). Dengue in the diabetic was characterized at presentation by more common cardiorespiratory signs and altered mental status but fewer muco-cutaneous signs.

Biologically, DPs had lower haematocrit and haemoglobin levels, but higher leucocyte counts than NDPs at presentation. However, the differences in white blood cell related parameters were substantial only for neutrophil count, the neutrophil to lymphocyte ratio (NLR), and the monocyte to platelet ratio (MPR), which higher values were potential indicators of dengue in the DP. Other biological parameters substantially associated with dengue in the DP include higher neutrophil to platelet ratio (NPR), neutrophil to lymphocyte*platelet ratio (NLPR), lipase, potassium, urea, and creatinine levels, while international normalized ratio, fibrinogen, sodium, chloride, urea to creatinine ratio, or C-reactive protein levels exhibited also significant differences in medians but not over the whole extent of their distributions. Noteworthy, there were no significant differences between the two groups for platelet counts and liver enzymes levels at presentation.

The abovementioned stringent selection procedure of covariates characterizing dengue in the DP identified four symptoms, three clinical features and four biological parameters, as potential indicators of dengue in the diabetic at first presentation (Table 2).

**Table 2.**
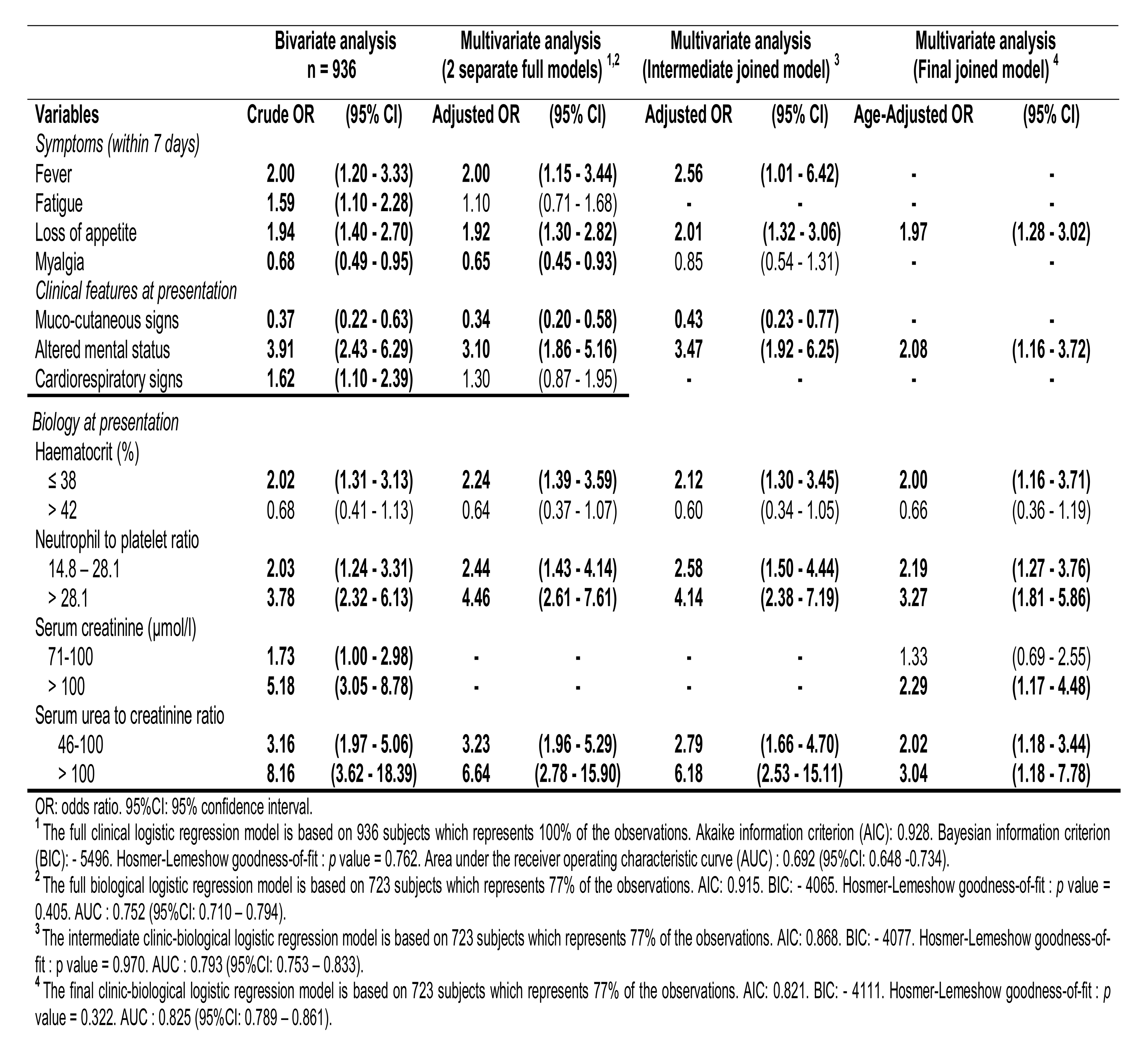
Clinical and biological characteristics found at hospital presentation in the diabetic patients with virologically confirmed cases of dengue, Reunion anuary to June 2019

| Variables | Bivariate analysis<br>n = 936 |  | Multivariate analysis<br>(2 separate full models) <sup>1,2</sup> |  | Multivariate analysis<br>(Intermediate joined model) <sup>3</sup> |  | Multivariate analysis<br>(Final joined model) <sup>4</sup> |  |
| --- | --- | --- | --- | --- | --- | --- | --- | --- |
|  | Crude OR | (95% CI) | Adjusted OR | (95% CI) | Adjusted OR | (95% CI) | Age-Adjusted OR | (95% CI) |
| *Symptoms (within 7 days)* |  |  |  |  |  |  |  |  |
| Fever | **2.00** | **(1.20 - 3.33)** | **2.00** | **(1.15 - 3.44)** | **2.56** | **(1.01 - 6.42)** | - | - |
| Fatigue | **1.59** | **(1.10 - 2.28)** | 1.10 | (0.71 - 1.68) | - | - | - | - |
| Loss of appetite | **1.94** | **(1.40 - 2.70)** | **1.92** | **(1.30 - 2.82)** | **2.01** | **(1.32 - 3.06)** | **1.97** | **(1.28 - 3.02)** |
| Myalgia | **0.68** | **(0.49 - 0.95)** | **0.65** | **(0.45 - 0.93)** | 0.85 | (0.54 - 1.31) | - | - |
| *Clinical features at presentation* |  |  |  |  |  |  |  |  |
| Muco-cutaneous signs | **0.37** | **(0.22 - 0.63)** | **0.34** | **(0.20 - 0.58)** | **0.43** | **(0.23 - 0.77)** | - | - |
| Altered mental status | **3.91** | **(2.43 - 6.29)** | **3.10** | **(1.86 - 5.16)** | **3.47** | **(1.92 - 6.25)** | **2.08** | **(1.16 - 3.72)** |
| Cardiorespiratory signs | **1.62** | **(1.10 - 2.39)** | 1.30 | (0.87 - 1.95) | - | - | - | - |
| *Biology at presentation* |  |  |  |  |  |  |  |  |
| Haematocrit (%) |  |  |  |  |  |  |  |  |
| ≤ 38 | **2.02** | **(1.31 - 3.13)** | **2.24** | **(1.39 - 3.59)** | **2.12** | **(1.30 - 3.45)** | **2.00** | **(1.16 - 3.71)** |
| > 42 | 0.68 | (0.41 - 1.13) | 0.64 | (0.37 - 1.07) | 0.60 | (0.34 - 1.05) | 0.66 | (0.36 - 1.19) |
| Neutrophil to platelet ratio |  |  |  |  |  |  |  |  |
| 14.8 – 28.1 | **2.03** | **(1.24 - 3.31)** | **2.44** | **(1.43 - 4.14)** | **2.58** | **(1.50 - 4.44)** | **2.19** | **(1.27 - 3.76)** |
| > 28.1 | **3.78** | **(2.32 - 6.13)** | **4.46** | **(2.61 - 7.61)** | **4.14** | **(2.38 - 7.19)** | **3.27** | **(1.81 - 5.86)** |
| Serum creatinine (μmol/l) |  |  |  |  |  |  |  |  |
| 71-100 | **1.73** | **(1.00 - 2.98)** | - | - | - | - | 1.33 | (0.69 - 2.55) |
| > 100 | **5.18** | **(3.05 - 8.78)** | - | - | - | - | **2.29** | **(1.17 - 4.48)** |
| Serum urea to creatinine ratio |  |  |  |  |  |  |  |  |
| 46-100 | **3.16** | **(1.97 - 5.06)** | **3.23** | **(1.96 - 5.29)** | **2.79** | **(1.66 - 4.70)** | **2.02** | **(1.18 - 3.44)** |
| > 100 | **8.16** | **(3.62 - 18.39)** | **6.64** | **(2.78 - 15.90)** | **6.18** | **(2.53 - 15.11)** | **3.04** | **(1.18 - 7.78)** |

OR: odds ratio. 95%CI: 95% confidence interval.
<sup>1</sup> The full clinical logistic regression model is based on 936 subjects which represents 100% of the observations. Akaike information criterion (AIC): 0.928. Bayesian information criterion (BIC): - 5496. Hosmer-Lemeshow goodness-of-fit : *p* value = 0.762. Area under the receiver operating characteristic curve (AUC) : 0.692 (95%CI: 0.648 -0.734).
<sup>2</sup> The full biological logistic regression model is based on 723 subjects which represents 77% of the observations. AIC: 0.915. BIC: - 4065. Hosmer-Lemeshow goodness-of-fit : *p* value = 0.405. AUC : 0.752 (95%CI: 0.710 – 0.794).
<sup>3</sup> The intermediate clinic-biological logistic regression model is based on 723 subjects which represents 77% of the observations. AIC: 0.868. BIC: - 4077. Hosmer-Lemeshow goodness-of-fit : *p* value = 0.970. AUC : 0.793 (95%CI: 0.753 – 0.833).
<sup>4</sup> The final clinic-biological logistic regression model is based on 723 subjects which represents 77% of the observations. AIC: 0.821. BIC: - 4111. Hosmer-Lemeshow goodness-of-fit : *p* value = 0.322. AUC : 0.825 (95%CI: 0.789 – 0.861).

Pre-existing chronic conditions and their related treatments were deemed indicative of diabetes comorbidities and not eligible to this analysis.

The three-step multivariate analysis identified fever, loss of appetite and altered mental status as independent clinical indicators, and low haematocrit (≤ 38%), high NPR ratio (≥ 14.8), upper-range creatinine (≥ 100 µmol/l) and high serum urea to creatinine (> 45) as independent biological indicators of dengue in the diabetic, whereas myalgia and muco-cutaneous signs were associated with dengue in the NDPs (data not shown).

In the pre-final joined model, adjusting for age abrogated the association between muco-cutaneous signs and NDPs (data not shown). The final age-adjusted joined model confirmed six independent indicators of dengue in the diabetic: loss of appetite (adjusted OR 1.97, 95%CI 1.28-3.02), altered mental status (aOR 2.02, 95%CI 1.28-3.02), low haematocrit (aOR 2.00, 95%CI 1.16-3.71), high NPR (aOR 2.19 in the intermediate tercile, aOR 3.27 in the upper tercile), upper-range creatinine (aOR 2.02, 95%CI 1.28-3.02) and high serum urea to creatinine ratio (aOR 2.02 in the intermediate tercile, aOR 3.04 in the upper tercile).

### Dengue in diabetic patients with or without diabetes complications

When compared to uncomplicated diabetes, dengue of the DPs with complicated diabetes occurred in older and highly comorbid subjects (S2 Table). At presentation, it was clinically unremarkable but characterized by higher serum creatinine level and slightly lower haematocrit, haemoglobin, or ALT levels. However, upon hospitalization, it was marked by higher peak C reactive protein and higher odds to evolve towards SD.

When compared to NDPs, DPs with diabetes complications exhibited more frequent dehydration, more clinical fluid accumulation and more haemoconcentration. Loss of appetite and altered mental status, indicators of inflammation or renal dysfunction were found in DPs, irrespective of the severity of diabetes.

The full abnormalities found given the severity of diabetes are displayed in S1 Fig.

### Dengue in diabetic patients with optimal or suboptimal glycaemic control

When compared to diabetes with optimal control (HBA1c <7%), dengue of the DP with suboptimal glycaemic control (HBA1c ≥ 7%) at presentation (S3 Table) was also clinically unremarkable but biologically characterized by higher MPR, lower sodium and chlorine levels.

When compared to NDPs, DPs with suboptimal glycaemic control expressed roughly the same particularities than DPs with clinically complicated diabetes, which showed a fairly overlap between clinical and biological diabetes disease markers.

The full abnormalities found given the control of diabetes are displayed in S2 Fig.

Taken together, these data suggest that thrombocytopenia (platelet counts < 100 G/l) in the diabetic patient was more likely indicative of an uncomplicated diabetes with suboptimal glycaemic control rather than a complicated diabetes.

### Predictors of severe dengue in the diabetic patients

The most important differences between DF (hereafter designated as non-severe dengue) and SD in the DPs are presented in Table 3, while the full comparison between both populations is displayed in S4 Table.

**Table 3.** Characteristics at hospital presentation of virologically confirmed cases of non severe and severe dengue cases among diabetic patients, Reunion island, January to June 2019.

| Variable | Total |  | Non severe dengue (DF) |  | Severe dengue (SD) |  | p value |
| --- | --- | --- | --- | --- | --- | --- | --- |
|  | n=184 | (%) | n=124 | (%) | n=60 | (%) |  |
| *Socio-demographic* |  |  |  |  |  |  |  |
| Age (years ; medians, Q<sub>1</sub>-Q<sub>3</sub>) | 70.0 | (59.4 - 77.7) | 70.0 | (58.6 - 77.3) | 69.6 | (60.6 - 78.4) | 0.875 |
| Female sex | 102 | (55.4) | 68 | (54.8) | 34 | (56.7) | 0.815 |
| *Comorbidities* |  |  |  |  |  |  |  |
| Chronic kidney disease | 40 | (21.7) | 21 | (16.9) | 19 | (31.7) | **0.023** |
| Charlson score (medians, Q<sub>1</sub>-Q<sub>3</sub>) | 5 | (3 - 7) | 4 | (3 - 6) | 6 | (3 - 8) | **0.018** |
| *Symptoms (within 7 days)* |  |  |  |  |  |  |  |
| Backache | 34 | (18.5) | 28 | (22.6) | 6 | (10.0) | **0.039** |
| *Clinical features at presentation* |  |  |  |  |  |  |  |
| Indicators of probable dengue |  |  |  |  |  |  |  |
| Body aches | 124 | (67.4) | 90 | (72.6) | 34 | (56.7) | **0.031** |
| Warning signs |  |  |  |  |  |  |  |
| 0 | 116 | (63.0) | 83 | (66.9) | 33 | (55.0) | 0.079 |
| 1 | 52 | (28.3) | 34 | (27.4) | 18 | (30.0) |  |
| > 1 | 16 | (8.7) | 7 | (5.7) | 9 | (15.0) |  |
| Clinical fluid accumulation (n=155) | 12 | (7.7) | 3 | (2.9) | 9 | (17.0) | **0.003** |
| Other manifestations |  |  |  |  |  |  |  |
| Dehydration | 69 | (37.5) | 40 | (32.3) | 29 | (48.3) | **0.035** |
| Cardiorespiratory signs | 45 | (24.5) | 18 | (14.5) | 27 | (45.0) | **< 0.001** |
| Altered mental status | 36 | (19.6) | 18 | (14.5) | 18 | (30.0) | **0.013** |
| Non severe bleeding | 21 | (11.4) | 9 | (7.3) | 12 | (20.0) | **0.011** |
| Cough | 17 | (9.2) | 7 | (5.6) | 10 | (16.7) | **0.016** |
| *Biology at presentation (medians, Q<sub>1</sub>-Q<sub>3</sub>)* |  |  |  |  |  |  |  |
| Neutrophil to lymphocyte ratio | 5.5 | (2.8 - 10.7) | 6.5 | (3.2 - 10.8) | 4.2 | (2.5 - 10.2) | **0.039** |
| AST (IU/l) | 42 | (24 - 79) | 37 | (22 - 70) | 46 | (31 - 103) | 0.070 |
| ALT (IU/l) | 26 | (16 - 52) | 23 | (14 - 43) | 37 | (20 - 66) | 0.216 |
| Alkaline phosphatase (IU/l) | 73 | (55 - 91) | 62 | (54 - 87) | 80 | (72 - 106) | **0.015** |
| Potassium (mmol/l) | 4.1 | (3.7 - 4.5) | 4.0 | (3.7 - 4.3) | 4.3 | (3.8 - 4.7) | **0.008** |

Data are numbers and column percentages, or medians and interquartile ranges (Q<sub>1</sub>-Q<sub>3</sub>) when specified. Percentages are compared using chi<sup>2</sup> or Fisher's Exact test, as appropriate. Medians are compared using a non-parametric Brown-Mood k-sample test and when it was significant, further with median and quartile regressions. \* p< 0.05, \*\* p<0.01 and \*\*\* p<0.001 for the less significant of both quantile regressions (no \* means that at least one quantile regression is not significant).

Age and gender distributions did not differ between DPs with SD and DPs with non-severe dengue.

DPs with SD were more frequently affected by chronic kidney disease (CKD, 31.7% *vs* 16.9%, p=0.023) and higher Charlson comorbidity index scores (median score 6; Q_1_-Q_3_: 3-8 *vs* 4, Q_1_-Q_3_: 3-6, *p*=0.018) than DPs with non-severe dengue.

Backache was less frequently reported in DPs with SD than in DPs with non-severe dengue. DPs with SD suffered less body aches but experienced more commonly non-severe bleeding, fluid accumulation, dehydration, cardiorespiratory signs (including cough) and altered mental status at presentation.

Biologically, DPs with SD exhibited lower median neutrophil to lymphocyte ratios but higher median phosphatase alkaline and potassium levels than DPs with non-severe dengue at presentation. However, these differences were not observed over the whole extent of variable distributions. Other biological parameters were similar between the two groups.

The selection procedure of covariates predictive of SD in the DPs identified diabetes complications (*i.e.*, diabetes with retinopathy and/or neuropathy and/or nephropathy and/or diabetic foot), CKD (especially when alleged of non-diabetic origin), untreated hypertension and entrenched-proxy Charlson comorbidity index *(i.e.*, Charlson score subtracted from diabetes-associated points) as potential comorbidities, non-severe bleeding, presence of warning signs, altered mental status or cough as potential early clinical features predictive of severe dengue in the DPs at presentation (Table 4).

**Table 4.** Anamnestic and clinical predictors of severe dengue at hospital presentation of a diabetic patient virologically confirmed dengue (RT-PCR+ or NS1 antigen+), Reunion island, January to June 2019

| Variables | Bivariate analysis |  | Multivariate analysis<br>(2 separate full models) <sup>1,2</sup> |  | Multivariate analysis<br>(1 final joined model) <sup>3</sup> |  |
| --- | --- | --- | --- | --- | --- | --- |
|  | Crude RR | (95% CI) | Adjusted RR | (95% CI) | Adjusted RR | (95% CI) |
| *Comorbidities* |  |  |  |  |  |  |
| Diabetes complications * | **1.84** | **(1.23 - 2.75)** | **1.74** | **(1.10 - 2.75)** | **2.06** | **(1.39 - 3.05)** |
| Chronic kidney disease of undifferentiated origin | **1.67** | **(1.10 - 2.53)** | - | - | - | - |
| Chronic kidney disease alleged of diabetic origin | 1.45 | (0.89 - 2.36) | - | - | - | - |
| Chronic kidney disease alleged of non-diabetic origin | 1.74 | (1.00 - 3.01) | 1.66 | (0.82 - 3.36) | - | - |
| Untreated hypertension | **1.85** | **(1.13 - 3.02)** | 1.43 | (0.84 - 2.41) |  |  |
| Entrenched-proxy Charlson comorbidity score ** | **1.09** | **(1.00 - 1.17)** | 1.01 | (0.90 - 1.13) |  |  |
| *Clinical features at presentation* |  |  |  |  |  |  |
| Warning signs *** |  |  |  |  |  |  |
| 1 | 1.22 | (0.75 - 1.95) | 1.25 | (0.75 - 2.07) | - | - |
| > 1 | **1.98** | **(1.17 - 3.33)** | 1.57 | (0.82 - 3.04) | - | - |
| Non severe bleeding | **1.94** | **(1.24 - 3.02)** | 1.65 | (0.91 - 2.95) | **2.34** | **(1.49 - 3.67)** |
| Altered mental status | **1.76** | **(1.16 - 2.67)** | **1.71** | **(1.15 - 2.54)** | **1.69** | **(1.15 - 2.50)** |
| Cough | **1.96** | **(1.23 - 3.12)** | **1.91** | **(1.18 - 3.07)** | **1.95** | **(1.25 - 3.06)** |

\* Diabetic retinopathy and/or diabetic neuropathy and/or diabetic nephropathy and/or diabetic foot.
\*\*The Charlson comorbidity score was entrenched (or reduced) of the points provided by diabetes and chronic kidney disease to allow the adjustment of the score with these factors without overfitting the model.
\*\*\* Abdominal pain or tenderness, persistent vomiting, clinical fluid accumulation, mucosal bleed, lethargy or restlessness, liver enlargement > 2 cm, increase in haematocrit concurrent with rapid decrease in platelet count.
RR: relative risk. 95%CI: 95% confidence interval.
<sup>1</sup> The full anamnestic Poisson regression model is based on 184 subjects which represents 100% of the observations. Akaike information criterion: 1.393. Bayesian information criterion: - 687. Area under the receiver operating characteristic curve (AUC) : 0.636 (95%CI: 0.547 - 0.725).
<sup>2</sup> The full clinical Poisson regression model is based on 184 subjects which represents 100% of the observations. AIC: 1.386. BIC: - 685. AUC : 0.675 (95%CI: 0.592 - 0.757).
<sup>3</sup> The final clinic-anamnestic Poisson regression model is based on 184 subjects which represents 100% of the observations. AIC: 1.343. BIC: - 696. AUC: 0.725 (95%CI: 0.648 - 0.802).

The final joined model identified four key independent predictors of severe dengue in the DPs: diabetes complications (adjusted RR 2.06, 95%CI 1.39-3.05), non-severe bleeding (aRR 2.34, 95%CI 1.49-3.67), altered mental status (aRR 1.69, 95%CI 1.15-2.50) and cough.

Among diabetes complications, diabetic retinopathy (16.7% vs 7.2%, p=0.049), and neuropathy (16.7% vs 5.5%, p=0.016) but not diabetic nephropathy (25.0% vs 14.5%, p=0.082) nor diabetic foot (3.1% vs 1.6%, p=0.597) were predictive of severe dengue.

## Discussion

To the best of our knowledge, this retrospective cohort study is the first that attempted to both characterize dengue and to identify predictors of severe dengue in the diabetic patient.

### Key findings

The first model identifies six independent indicators of dengue in the DPs, namely loss of appetite, altered mental status, low haematocrit (≤ 38%), high neutrophil to platelet ratio (≥ 14.8), upper-range creatinine (> 100 µmol/l), and high serum urea to creatinine ratio (> 45). To a lesser extent, this model also showed that muco-cutaneous signs were indicative of dengue in the NDPs, but the association did not resist to age adjustment.

The second model proposes four key predictors of SD in the DPs, namely diabetes complications, non-severe bleeding, altered mental status and cough.

### Strengths and limitations

This study has several strengths. First, the cohort was extracted from the EPIDENGUE database, a quasi-exhaustive register of all dengue cases having consulted at the University hospital in 2019, which allows our study population to be considered as representative of a hospital dengue-infected population in Reunion. Second, the cohort was recruited during a single dengue seasonal epidemic in the hospital setting, which ensures, theoretically, a relative homogeneity in the definition and classification of dengue cases, especially as the Reunionese population was naïve to dengue on the study period. Third, with respect to the topic, the study sample was relatively important, and the selection method of candidate indicators/predictors was rigorous, which allowed the possibility of conclusive multivariate models on both aspects of the analysis. Fourth, the two final multivariate models displayed acceptable metrics both in terms of fitness, calibration, and discrimination, which confers internal validity to the analysis.

This study has also some limitations. First, given the retrospective character of this cohort study, the study may be prone to reporting bias (i.e., mainly underreporting of exposure/events), which may have hampered the completeness of the data and be the source of subsequent information or evaluation biases, this depending on whether it is an exposure or an outcome variable which is affected. Second, this hospital-based study might have been also submitted to a potential selection or referral bias, given the only hospital recruitment of dengue cases. However, we believe this do not really stand for most of the DPs, diabetes being recognized as a risk factor of severe infection for most emerging infectious diseases [19-21]. This may have been incentive for the primary care physician to refer the DPs of suspected dengue to the hospital, as suggested by high hospitalization rates including for DPs with uncomplicated diabetes.

### Interpretation

Diabetic patients represented 19.6% of the study population. This proportion overlaps more or less the figure expected in the Reunion island adult population, given former estimates of the prevalence of diabetes at 20.1% in the Reunionese aged 30 to 69 [22]. DPs were older and more comorbid than NDPs, which may illustrate the fact that diabetes is an age-related disease that along with aging tends to cluster other chronic conditions. This element might have skewed the DPs towards higher proportions of complicated diabetes as compared to the overall population of diabetics observed on the island. In addition, some DPs may have been referred to the hospital by their primary care physician because they had comorbidities or warning signs, or need to upgrade the antidiabetic medication (*e.g.,* insulin in support to oral treatment).

DPs with dengue were more likely to present loss of appetite than NDPs, which could suggest the presence of subclinical exocrine pancreatitis in some patients [23]. Consistent with this, we found among DPs with loss of appetite, higher lipase levels, more common nausea/vomiting, and non-significant trends towards more persistent vomiting or abdominal tenderness than in DPs without anorexia (data not shown). Whether this may be attributable to dengue or diabetes, or both, cannot be assessed from our data.

DPs with dengue exhibited more commonly altered mental status (*i.e.,* confusion, behavioural changes, or other cognitive impairments), the major criterion for defining encephalitis [24]. This finding was unrelated to several potential indicators of plasma leakage, endothelial activation or systemic inflammation in stratified analysis (*e.g.*, haemoconcentration, fluid accumulation, NLR, PLR) but coherent with that observed in West Nile flavivirus encephalitide, for which diabetes is a known risk factor [25].

DPs with dengue had lower haematocrit than NDPs, which correlated with lower haemoglobin and higher C reactive protein levels and was potentiated by CKD. Together, this finding would better account for inflammatory anaemia due to diabetes (especially if complicated with nephropathy) rather than for a proper effect of dengue [26]. Alternatively, DPs with complicated diabetes exhibited also more haemoconcentration than NPDs, which raises the interest in new biomarkers of fluid accumulation than the haematocrit alone (or the haematocrit plus the rapid platelet count fall) to better assess the plasma leakage in this population.

Interestingly, DPs with dengue displayed a higher NPR, a predictor that has been linked to dengue mortality or need for critical care in our setting [27]. To the best of our knowledge, this ratio had not been used previously in dengue neither for diagnosis nor for prognosis purpose. Given it was correlated both to NLPR and NLR (data not shown), we hypothesize that it could be indicative of both systemic inflammation [28] and early platelet-neutrophil interactions aimed at clearing the virus through neutrophil extracellular traps (NETs), this latter mechanism to be triggered by activated platelets in dengue [29] but also well-known to be at work in diabetes through a different pathway [30]. In addition, a high NPR at hospital presentation was later associated with thrombocytopenia and lower platelet nadirs when the full hospital stay was considered (data not shown), which may support shared, or even additive, mechanisms in both diabetic and SD patients in the pathogenesis of thrombocytopenia.

Dengue in the DPs was independently associated with both organic and functional components of acute kidney injury, as suggested by higher serum urea to creatinine ratio and higher serum creatinine level than in NDPs. However, subsequent stratified analyses (data not shown) revealed that in DENV-infected DPs with CKD, only the organic component was prevailing, which contrasted with NDPs with CKD for which the two components contributed to DENV-driven deterioration in renal function. Taken together, these data may illustrate the multifactorial mechanism of dengue-related acute kidney injury [31] and the greater sensitivity of the diabetic kidney to DENV infection [32].

Finally, this study proposes the presence of diabetes complications as an independent predictor of SD for the DP from a model controlling the other comorbidities, warning signs, and three early predictors of SD. Importantly, the model reveals that diabetes supplants all other comorbidities in the development of SD, especially hypertension and CKD, two other highly prevalent chronic conditions on the island [33, 34]. It also suggests that DPs with complicated diabetes are prone to all three modes of entry into SD, namely severe bleeding, severe plasma leakage with respiratory distress, and severe central nervous system involvement, with non-severe bleeding, cough and altered mental status, as respective harbingers. Consistent with this hypothesis, these harbingers were associated with more severe related organ dysfunctions in subsequent analyses (*e.g.*, cough with acute lung injury, data not shown).

Among the mechanisms raised to promote the aggravation of dengue in the DP, apoptosis of leucocytes and endothelial microvascular cells has been proposed from post-mortem findings [35], while more recently, a mouse model unravelled the role of hyperglycaemic stress to facilitate DENV replication through the poly(A)-binding protein PI3K/AKT signalling [36]. Together with the microvascular and macrovascular damages of diabetes complications, this would increase the susceptibility of the DPs to the severe plasma leakage key to SD pathogenesis [37]. In our study, the lack of correlates within the subgroups of complicated diabetes and DPs with known HbA1c levels deters to propose a putative mechanism. Nevertheless, we assume that the higher MPR found in DPs with HbA1c ≥ 7% than that observed in DPs with HbA1c <7% might reflect more intense trafficking of monocytes under monocyte chemoattractant protein-1 secretion induced by activated platelets [35, 38], which might also increase endothelial dysfunction and support more severe plasma leakage in the DP with suboptimal control [39].

### Generalizability

Notwithstanding the monocentric nature of this hospital-based study, several results echo observations published in the literature and suggest that some of its findings could be extrapolated, at least to other DENV-2 epidemics in similar contexts.

First, we confirm that diabetes outweighs the other comorbidities in promoting the pathogenesis of SD, which is consistent with the meta-analysis by Sangkaew et al. that showed the highest pooled OR for diabetes in the progression from DF to SD in random-effects models controlling prognostic factors [40]. Moreover, our findings support the importance of diabetes control aimed at preventing its vascular complications to avoid SD. Together, these elements corroborate the results of Pang et al. in Singapore and of Lee et al. in Taiwan who have shown, both in populations mainly affected by DENV-2 that only DPs with additional comorbidities, especially hypertension, were at increased risk of severe forms of dengue [13, 39].

Second, the study confirms the importance of non-severe bleeding disorders in the DPs as a predictor of SD, which is consistent with studies identifying diabetes as a risk factor for dengue haemorrhagic fever (WHO 1997 classification) [13] or a risk factor for SD (WHO 2009 classification) [39]. Surprisingly, we did not find lower admission platelet counts in DPs than in NDPs, which differs from the recent observation by Singh et al. in India [37] and another one in Taiwan. However, on Reunion island, DPs exhibited lower platelet nadir than NDPs upon hospitalization, which may relate the discrepancy between studies to the earlier presentation in the course of illness of DPs (75% within 3 days post-symptom onset). Indeed, platelet nadir during dengue usually occurs within 4-5 days (48 hours of the critical phase) and our observation of a platelet nadir at day 2 in DPs lends to support to this explanation.

Third, we identified altered mental status, as a predictor of SD in the DPs, which is consistent with the increased odds of intracranial haemorrhage or cerebral infarction found among Taiwanese patients with dengue and altered consciousness in multivariate analyses controlling potential comorbidities [41]. Interestingly, DPs represented a third of their study sample, and one model adjusted for a Charlson score >4, which matches 60% of DPs and 96% of DPs with complicated diabetes in our study. Beyond the mode of entry into dengue encephalitis, this raises the possibility that our second model may apply to predict severe central nervous involvement, as results of haemorrhagic or thrombotic events, more frequent in the DPs.

Fourth, we identified cough as a predictor of SD in the DPs. Although respiratory tract involvement is uncommon in dengue, cough has been reported in approximately 25–50% of DF cases. This result is consistent with the Thai report by Temprasertrudee, which identified cough as an independent predictor of SD in a model controlling alanine aminotransferase levels [42].

Last, the findings of the study evidences a close relationship between diabetes and renal dysfunction among dengue patients which may support diabetes as risk factor for acute dengue-related kidney injury [32], even though such a study remains to be done in our context.

### Implications

For individual health purposes, this study may help patient management through better characterization of dengue and identification of early prognostic factors of SD in the DPs. Indeed, this should encourage emergency physicians during epidemic periods to refine the triage of patients consulting the emergency department and propose more intense monitoring of DPs with diabetes complications to allow earlier hospitalization whenever needed.

For public health purposes, this study should motivate more active screening strategies of both conditions among people living in or travelling to dengue endemic countries and emphasize the DPs with complicated diabetes as a candidate for dengue vaccine immunization.

For research purposes, this study should help to design future studies aimed at investigating the interactions between diabetes and dengue and the concept of syndemic.

There are several arguments for a double negative impact of both affections each on the other and understanding the short-term and long-term consequences of dengue in the DP deserves certainly further studies.

## Conclusion

At hospital first presentation, dengue in the diabetic patient is characterized by deteriorations in appetite, mental and renal functioning, while severe dengue can be predicted by diabetes complications, dengue-related non-severe haemorrhages, cough, and dengue-related encephalopathy. Larger prospective studies are needed to confirm these findings and decipher their pathogenic significance.

## Data availability statement

All data produced in the present study are available upon reasonable request to the authors.

## Credit authorship contribution

**Azizah Issop**: Investigation, Interpretation of the data, Original draft writing - review and editing. **Antoine Bertolotti**: Formal analysis, Supervision - review and editing. **Yves-Marie Diarra**: Formal analysis, Methodology - review and editing. **Jean-christophe Maïza**: Investigation - review and editing. **Eric Jarlet**: Investigation - review and editing. **Muriel Cogne**: Investigation - review and editing. **Eric Doussiet**: Investigation - review and editing. **Eric Magny**: Investigation, Software - review and editing. **Olivier Maillard**: Formal analysis, Project administration - review and editing. **Epidengue Cohort Investigation Team**: Jeanne Belot, Mathilde Cadic, Mathys Carras, Romain Chane-Teng, Romane Crouzet, David Hirschinger, Anne-Cecilia Etoa N’Doko, Azizah Issop, Mathilde Legros, Mamitiana Randriamanana, Cédric Rosolen, Nolwenn Sautereau, Investigation – review and editing. **Estelle Nobécourt**: Formal analysis, Supervision, Writing - review and editing. **Patrick Gérardin**: Conceptualization, Data curation, Formal analysis, Methodology, Software, Validation, Vizualisation, Writing – review and editing.

## Supporting information

STROBE Checklist

## Data Availability

All data produced in the present study are available upon reasonable request to the authors

## Acknowledgments

The authors thank the emergency units of the University Hospital of Reunion and all collaborators of the EPIDENGUE project.

## References

1) Du M, Jing W, Liu M, Liu J. The global trends and regional differences in incidence of dengue infection from 1990 to 2019: an analysis from the Global Burden of Diseases of Study. Infect Dis Ther 2021; 10: 1625–43. https://dx.doi/10.1007/s40121-021-00470-2.

2) European Center for Disease Prevention and Control. Dengue worldwide overview. https://www.ecdc.europa.eu/en/dengue-monthly. [Accessed 4 April 2023].

3) Semenza JC, Rocklöv J, Ebi KL. Climate change and cascading risks from infectious disease. Infect Dis Ther. 2022; 11:1371–90. https://dx.doi/10.1007/s40121-022-00647-3.

4) La Ruche G, Souares Y, Armengaud A, Peloux-Petiot F, Delaunay P, Despres P, et al. First two autochthonous dengue virus infections in metropolitan France, September 2010. Euro Surveill 2010; 15: 19676. https://dx.doi/10.2807/ese.15.39.19676-en.

5) Cochet A, Calba C, Jourdain F, Grard G, Durand GA, Guinard A, et al. Autochthonous dengue in mainland France, 2022: geographical extension and incidence increase. Euro Surveill. 2022; 27: 2200818. https://dx.doi/10.2807/1560-7917.ES.2022.27.44.2200818.

6) Vanwambecke SO, Bennett SN, Kapan DD. Spatially disaggregated disease transmission risk: land use, land cover and risk of dengue transmission on the island of Ohau Trop Med Int Health 2011; 16: 174-85. https://dx.doi/10.1111.j1365-3156.2010.02671.x

7) Santé Publique France. DENGUE. Point epidemiologique hebdomadaire, La Reunion, 7 decembre 2021. Sante publique France-Reunion 6 pp. https://www.santepubliquefrance.fr/regions/ocean-indien/documents/bulletin-regional/2021/surveillance-de-la-dengue-a-la-reunion.-point-au-7-decembre-2021. [Accessed 4 April 2023].

8) World Health Organization. Dengue guidelines for diagnosis, treatment, prevention and control□: new edition. 2009. https://apps.who.int/iris/handle/10665/44188. [Accessed 4 April 2023].

9) Joshi N, Caputo GM, Weitekamp MR, Karchmer AW. Infections in patients with diabetes mellitus. N Engl J Med. 1999; 341: 12. https://dx.doi/10.1056/NEJM199912163412507.

10) Bertoni AG, Saydah S, Brancati FL. Diabetes and the risk of infection-related mortality in the U.S. Diabetes Care. 2001; 24: 1044–9. https://dx.doi/10.2337/diacare.24.6.1044.

11) Shah BR, Hux JE. Quantifying the risk of infectious diseases for people with diabetes. Diabetes Care. 2003;26: 510–3. https://dx.doi/10.2337/diacare.26.2.510.

12) Htun NSN, Odermatt P, Eze IC, Boillat-Blanco N, D’Acremont V, Probst-Hensch N. Is diabetes a risk factor for a severe clinical presentation of dengue? - Review and meta-analysis. PLoS Negl Trop Dis. 2015; 9: e0003741. https://dx.doi/10.1371/journal.pntd.0003741.

13) Pang J, Salim A, Lee VJ, Hibberd ML, Chia KS, Leo YS, et al. Diabetes with hypertension as risk factors for adult dengue hemorrhagic fever in a predominantly dengue serotype 2 epidemic: a case control study. PLoS Negl Trop Dis. 2012; 6: e1641. https://dx.doi/10.1371/journal.pntd.0001641.

14) ORS La Réunion. ors_chiffres_diabete_reunion_2021.pdf. 2021. https://www.ors-reunion.fr/IMG/pdf/ors_chiffres_diabete_reunion_2021.pdf.

15) Brown GW, Mood AM. On median tests for linear hypotheses. In: Proceeding of the second Berkeley symposium on mathematical statistics and probability, University of California Press ed, Berkeley, 1951: pp 159–66.

16) Chamberlain G. Quantile regression, censoring, and the structure of wages. In: Advances in Economics Sixth World Congress, Cambridge University Press Sims CA ed, Cambridge 1994: pp 171-209.

17) Zou G. A modified poisson regression approach to prospective studies with binary data. Am J Epidemiol 2004; 159(7): 702□6. https://dx.doi/10.1093/AJE/KWH090.

18) Riley RD, Ensor J, Snell KIE, Harrell Jr FE, Martin GP, Reitsma JB, et al. Calculating the sample size required for developing a clinical prediction model. BMJ 2020; 368:m441 https://dx.doi/10.1136/bmj.m441.

19) Badawi A, Ryoo SG, Vasileva D, Yaghoubi S. Prevalence of comorbidities in Chikungunya: a systematic review and meta-analysis. Int J Infect Dis. 2018; 67: 107–13. https://dx.doi/10.1016/j.ijid.2017.12.018 1201-9.

20) Badawi A, Ryoo SG, Vasileva D, Yaghoubi S. Prevalence of comorbidities in dengue fever and West Nile virus: a systematic review and meta-analysis. PLoS One 2018; 13: e0200200. https://dx.doi/10.1371/journal.pone.0200200.

21) Longmore DK, Miller JE, Bekkering S, Saner C, Mifsud E, Zhu Y et al. Diabetes and overweight/obesity are independent, nonadditive risk factors for in-hospital severity of COVID-19: an international, multicenter, retrospective meta-analysis. Diabetes Care 2021; 44: 1281–90. https://dx.doi/10.2337/dc20-2676.

22) Favier F, Jaussent I, Moullec NL, Debussche X, Boyer M-C, Schwager J-C, et al. Prevalence of Type 2 diabetes and central adiposity in La Réunion Island, the REDIA Study. Diabetes Res Clin Pract. 2005;67: 234–242. https://dx.doi/10.1016/j.diabres.2004.07.013.

23) Correa R, Ortega-Loubon C, Zapata-Castro LE, Armien B, Culquichicon C. Dengue with hemorhagic and acute pancreatitis: case report and review. Cureus 2019; 11: e4895. https://dx.doi/10.7759/cureus.4895.

24) Venkatesan A, Tunkel AR, Bloch KC, Lauring AS, Sejvar J, Bitnun A, et al. Case definitions, diagnostic algorithms, and priorities in encephalitis: consensus statement of the International Encephalitis Consortium. Clin Infect Dis 2013; 57: 1114–28. https://dx.doi/10.1093/cid/cit458.

25) Jean CM, Honarmand S, Louie JK, Glaser CA. Risk factors for West Nile virus neuroinvasive disease, California, 2005. Emerg Infect Dis 2007; 13:1918-20. https://dx.doi/10.3201/eid1312.061265.

26) Mehdi U, Toto RD. Anemia, diabetes, and chronic kidney disease. Diabetes Care 2009; 32: 1320–6. https://dx.doi/10.2337/dc08-0779.

27) Romane Crouzet. [Facteurs de risque de décès ou de thérapeutique invasive chez les patients suspects de dengue durant l’épidémie à La Réunion en 2019]. In: HAL-Réunion. Archive ouverte de l’Université de La Réunion, ed. Sciences du Vivant [q-bio] Dumas-03831531, v1. 2022, 79 pp. https://hal.univ-reunion.fr/dumas-03831531v1

28) Osuna-Ramos JF, Reyes-Ruiz JM, Ochoa-Ramírez LA, De Jesús-González LA, Ramos-Payán R, Farfan-Morales CN. The usefulness of peripheral blood cell counts to distinguish COVID-19 from dengue during acute infection. Trop Med Infect Dis 2022; 7: 20. https://dx.doi/10.3390/tropicalmed7020020.

29) Garisha F, Rother N, Riswari SF, Alisjabaah B, Overheul GJ, van Rij RP, et al. Neutrophil extracellular traps in dengue are mainly generated NOX-independently. Front Immunol 2021; 12: 629167. https://dx.doi/10.3389/fimmu.2021.629167.

30) Njeim R, Azar WS, Fares AH, Azar ST, Kfoury Kassouf H, Eid AA. NETosis contributes to the pathogenesis of diabetes and its complications. J Mol Endocrinol 2020; 65: R65–R76. https://dx.doi/10.1530/JME-20-0128.

31) Gurugama P, Jayarajah U, Wanigasuriya K, Wijewickrama A, Perera J, Seneviratne SL. Renal manifestations of dengue virus infections. J Clin Virol. 2018; 101: 1–6. https://dx.doi/10.1016/j.jcv.2018.01.001.

32) Mallhi TH, Khan AH, Adnan AS, Sarriff A, Khan YH, Jummaat F. Incidence, characteristics and risk factors of acute kidney injury among dengue patients: a retrospective analysis. PLoS One 2015; 10: e0138465. https://dx.doi/10.1371/journal.pone.0138465.

33) Stengel B, Jaussent I, Guiserix J, Bourgeon B, Papoz L, Favier F. High prevalence of chronic kidney disease in La Réunion island and its association with the metabolic syndrome in the non-diabetic population: La Réunion Diabetes (REDIA) Study. Diabetes Metab. 2007; 33: 444–52. https://dx.doi/10.1016/j.diabet.2007.10.002.

34) Cournot M, Lenclume V, Le Moullec N, Debussche X, Doussiet E, Fagot-Campagna A, Favier F. Prevalence, treatment, and control of hypertension in La Réunion: the REDIA population-based cohort study. Blood Press 2017; 26: 39–47. https://dx.doi/10.1080/08037051.2016.1182854.

35) Limonta D, Torres G, Capó V, Guzmán MG. Apoptosis, vascular leakage and increased risk of severe dengue in a type 2 diabetes mellitus patient. Diab Vasc Dis Res. 2008; 5: 213–4. https://dx.doi/10.3132/dvdr.2008.034.

36) Shen TJ, Chen CL, Tsai TT, Jhan MK, Bai CH, Yen YC, et al. Hyperglycemia exacerbates dengue virus infection by facilitating poly(A)-binding protein-mediated viral translation. JCI Insight 2022; 7: e142805.

37) https://dx.doi/10.1172/jci.insight.142805.

37) Singh R, Goyal S, Aggarwal N, Mehta S, Kumari P, SinghV, et al. Study on dengue severity in diabetic and non-diabetic population of tertiary care hospital by assessing inflammatory indicators. Ann Med Surg (Lond) 2022; 82: 104710. https://dx.doi/10.1016/j.amsu.2022.104710.

38) Gawaz M, Neumann FJ, Dickfeld T, Koch W, Laugwitz KL, Adelsberger H, et al. Activated platelets induce monocyte chemotactic protein-1 secretion and surface expression of intercellular adhesion molecule-1 on endothelial cells. Circulation 1998; 98: 1164–71. https://dx.doi/10.1161/01.cir.98.12.1164.

39) Lee IK, Hsieh C-J, Lee CT, Liu JW. Diabetic patients suffering dengue are at risk for development of dengue shock syndrome/severe dengue: Emphasizing the impacts of co-existing comorbidity(ies) and glycemic control on dengue severity. J Microbiol Immunol Infect. 2020; 53: 69–78. https://dx.doi/10.1016/j.jmii.2017.12.005.

40) Sangkaew S, Ming D, Boonyasiri A, Honeyford K, Kalayanarooj S, Yacoub S, et al. Risk predictors of progresion to severe diseases during the febrile phase of dengue: a systematic review and meta-analysis. Lancet Infect Dis 2021; 21: 1014–26. https://dx.doi/10.1016/S1473-3099(20)30601-0.

41) Chang K, Huang CH, Chen TC, Lin CY, Lu PL, Chen YH. Clinical characteristics and risk factors for intracranial haemorrhage or infarction in patients with dengue. J Microbiol Immunol Infect 2021; 54: 885–92. https://dx.doi/10.1016/j.jmii.2021.03.009.

42) Temprasertrudee S, Thanachartwet V, Desakorn V, Keatkla J, Chantratita W, Kiertinuranakul S. A multicenter study of clinical presentations and predictive factors for severe manifestations of dengue in adults. Jpn J Infect Dis 2018; 71: 239–43. https://dx.doi/10.7883/yoken.JJID.2017.457.

