## Supplementary material for "Dengue clinical features and predictors of severity in the diabetic patient: a retrospective cohort study on Reunion island, 2019": STROBE Checklist

STROBE Statement—checklist of items that should be included in reports of observational studies

|  | Item No. | Recommendation | Page  No. | Relevant text from manuscript |
| --- | --- | --- | --- | --- |
| **Title and abstract** | 1 | (*a*) Indicate the study’s design with a commonly used term in the title or the abstract | 1 | Retrospective cohort study |
|  |  | (*b*) Provide in the abstract an informative and balanced summary of what was done and what was found | 2 |  |
| Introduction | | | |  |
| Background/rationale | 2 | Explain the scientific background and rationale for the investigation being reported | 3 | Lines 62 to 86 |
| Objectives | 3 | State specific objectives, including any prespecified hypotheses | 3 | Lines 87 to 92 |
| Methods | | | |  |
| Study design | 4 | Present key elements of study design early in the paper | 4 | Lines 95 to 97 |
| Setting | 5 | Describe the setting, locations, and relevant dates, including periods of recruitment, exposure, follow-up, and data collection | 4 | Lines 95 to 117 |
| Participants | 6 | (*a*) *Cohort study*—Give the eligibility criteria, and the sources and methods of selection of participants. Describe methods of follow-up | 4 | Lines 95 to 111 |
| Variables | 7 | Clearly define all outcomes, exposures, predictors, potential confounders, and effect modifiers. Give diagnostic criteria, if applicable | 4-5 | Lines 101 to 156 |
| Data sources/ measurement | 8* | For each variable of interest, give sources of data and details of methods of assessment (measurement). Describe comparability of assessment methods if there is more than one group | 4-5 | Lines 120 to 156 |
| Bias | 9 | Describe any efforts to address potential sources of bias | 5 | Lines 140 to 156  confounding |
| Study size | 10 | Explain how the study size was arrived at | N.A | No prespecified calculation |

Continued on next page

| Quantitative variables | 11 | Explain how quantitative variables were handled in the analyses. If applicable, describe which groupings were chosen and why | 4-5 | Lines 124 to 127 |
| --- | --- | --- | --- | --- |
| Statistical methods | 12 | (*a*) Describe all statistical methods, including those used to control for confounding | 5 | Lines 130 to 148 |
|  |  | (*b*) Describe any methods used to examine subgroups and interactions | 5 | Lines 149 to 151 |
|  |  | (*c*) Explain how missing data were addressed | 5 | Lines 155 to 156 |
|  |  | (*d*) *Cohort study*—If applicable, explain how loss to follow-up was addressed | N.A |  |
|  |  | (*e*) Describe any sensitivity analyses | N.A |  |
| Results | | | | |
| Participants | 13* | (a) Report numbers of individuals at each stage of study—eg numbers potentially eligible, examined for eligibility, confirmed eligible, included in the study, completing follow-up, and analysed | 6 | Lines 170 to 175 |
|  |  | (b) Give reasons for non-participation at each stage | 6 | Lines 170 to 175 |
|  |  | (c) Consider use of a flow diagram | 6 | Line 173: Figure 1 |
| Descriptive data | 14* | (a) Give characteristics of study participants (eg demographic, clinical, social) and information on exposures and potential confounders | 7 | Lines 169 to 186,  Table 1, Table S1 |
|  |  | (b) Indicate number of participants with missing data for each variable of interest | N.D | Not done but when effective is drastically small, numbers are given to show how % are estimated  (ex: clinical fluid accumulation) |
|  |  | (c) *Cohort study*—Summarise follow-up time (eg, average and total amount) | N.A | This is not a longitudinal cohort with follow-up |
| Outcome data | 15* | *Cohort study*—Report numbers of outcome events or summary measures over time |  | Table 1, Table 3, Table S1 to S4 |
| Main results | 16 | (*a*) Give unadjusted estimates and, if applicable, confounder-adjusted estimates and their precision (eg, 95% confidence interval). Make clear which confounders were adjusted for and why they were included | 10,15 | Table 2, Table 4 |
|  |  | (*b*) Report category boundaries when continuous variables were categorized | 8-9,13 | Continuous variables are expressed as median with interquartile ranges in Table 1, Table 3, Table S1 to S4 |
|  |  | (*c*) If relevant, consider translating estimates of relative risk into absolute risk for a meaningful time period | N.A | There is no follow-up. Absolute risk are not given |

Continued on next page

| Other analyses | 17 | Report other analyses done—eg analyses of subgroups and interactions, and sensitivity analyses | 11 | Lines 226 to 242. Subgroup analyses for DPs are given Table S2 to Table S4 |
| --- | --- | --- | --- | --- |
| Discussion | | | | |
| Key results | 18 | Summarise key results with reference to study objectives | 17 | Lines 286 to 292 |
| Limitations | 19 | Discuss limitations of the study, taking into account sources of potential bias or imprecision. Discuss both direction and magnitude of any potential bias | 17,18 | Lines 307 to 317 |
| Interpretation | 20 | Give a cautious overall interpretation of results considering objectives, limitations, multiplicity of analyses, results from similar studies, and other relevant evidence | 18-20 | Lines 319 to 389 |
| Generalisability | 21 | Discuss the generalisability (external validity) of the study results | 20,21 | Lines 391 to 431 |
| Other information | |  | | |
| Funding | 22 | Give the source of funding and the role of the funders for the present study and, if applicable, for the original study on which the present article is based | 23 | Lines 455 to 458 |

*Give information separately for cases and controls in case-control studies and, if applicable, for exposed and unexposed groups in cohort and cross-sectional studies.

**Note:** An Explanation and Elaboration article discusses each checklist item and gives methodological background and published examples of transparent reporting. The STROBE checklist is best used in conjunction with this article (freely available on the Web sites of PLoS Medicine at http://www.plosmedicine.org/, Annals of Internal Medicine at http://www.annals.org/, and Epidemiology at http://www.epidem.com/). Information on the STROBE Initiative is available at www.strobe-statement.org.
