## Supplemental tables and figures for "Dengue clinical features and predictors of severity in the diabetic patient: a retrospective cohort study on Reunion island, 2019"

**Supplementary Files**

| **S0 Table. Definitions of dengue warning signs, severe dengue criteria, severe organ involvement and other non-severe manifestations of dengue, Reunion island, January to June 2019.** | |
| --- | --- |
| ***Warning signs*** |  |
| Abdominal pain or tenderness | Pain or tenderness of abdomen upon palpation |
| Persistent vomiting | Recurrent episodes of vomiting prior and after presentation to the hospital |
| Clinical fluid accumulation | Haemoconcentration (haematocrit increased > 15% of the normal value for age, sex and population, or a haematocrit decreased < 15% after vascular filling), or serous effusions or hypoalbuminemia < 25 g/l |
| Mucosal bleed | Nose, gum or conjunctival bleeding, macroscopic haematuria, unexpected abundant menorrhagia, metrorrhagia, melaena, rectorrhagia |
| Lethargy, restlessness | Prostration with drowsiness or agitation |
| Hepatomegaly | Palpable liver enlargement > 2 cm below costal margin |
| Increase in haematocrit concurrent with rapid decrease in platelet count | Haematocrit increased > 15% with concomitant decrease of platelet count < 50% of first measured platelet count or with thrombocytopenia < 50 G/l. |
| ***Severe dengue*** | **Severe plasma leakage leading to :** |
| Shock | Compensated: Rapid pulse for age and narrow differential blood pressure (< 20 mm); Decompensated: Hypotension and capillary hypoperfusion |
| Severe fluid accumulation | Fluid accumulation as defined above plus respiratory distress (SaO2 < 95% or need for oxygen supply) or signs of shock |
| Severe bleeding | Intracranial haemorrhage, alveolar haemorrhage, haemoptysis, upper or lower gut bleeding with need for platetet or red blood cell infusion |
|  | **Or severe organ involvement** : |
| Cardiac | Acute pulmonary oedema or cardiogenic shock or left ventricular shortening fraction < 50% or raised troponin I or raised troponin T |
| Central Nervous system | Glasgow coma score < 11 or Blantyre coma score < 3, or altered mental status plus ≥ 2 minor encephalitis criteria (as defined within IEC *) |
| Circulatory | Hypotension : systolic blood pressure < 80 mmHg (< 5 years) or < 90 mmHg (≥ 5 years) or mean blood pressure < 65 mmHg or need for pressor amines |
| Disseminated intravascular coagulation | D Dimer > 500 pg/l and (one of the following: platelet count < 50 G/l or prothrombin time < 50% or index normalized ratio >2) or two of the following: 50 G/l< platelet count <100 G/l or 50%< PT <65% or 1.5< INR <2. |
| Hepatic | AST or ALT > 1000 IU/l or jaundice with prothrombin time < 50 % |
| Renal | Serum creatinine > 354 µmol/l or increase in serum creatinine > 50% or need for renal replacement therapy (hemofiltration or haemodialysis) |
| Respiratory | Respiratory distress with PaO_2_/FiO_2_ < 300 or SaO2 < 95% and respiratory rate > 24 cycles per minute and need for oxygen supply or positive end-expiratory pressure |
| **Other manifestations** |  |
| Altered mental status | Lethargy, restlessness, personality (or behavioural) changes, with less than two minor additional criteria for encephalitis (as defined within IEC *) |
| Cardiorespiratory signs | Dyspnea (shortness of breath, at rest or on exertion), wheezing, chest pain, cough, sputum, palpitations, crackles or sub-crackles, or pleural effusion |
| Dehydration | Mucosal dryness, skin fold with tachycardia and need for intravenous rehydration |
| Non severe bleeding | Mucosal bleed as abovementioned or any type of purpura |
| ** IEC: International Encephalitis consortium* | |

| **S1 Table. Characteristics at hospital presentation of virologically confirmed cases of dengue among non-diabetic and diabetic patients, Reunion island, January to June 2019.** | | | | | | | | | | | |
| --- | --- | --- | --- | --- | --- | --- | --- | --- | --- | --- | --- |
| **Variable** | | **Total** | | **Non-diabetic patients (NDPs)** | | | **Diabetic patients (DPs)** | | | |  |
|  | n=936 | | (%) | n=752 | (%) | | n=184 | | | (%) | *p value* |
| *Socio-demographic* |  | |  |  |  | |  | |  | |  |
| Age (years ; medians, Q_1_-Q_3_) | 51.9 | | (32.7 - 69.7) | 45.6 | (29.6 - 63.1) | | 70.0 | | (59.4- 77.7) | | *****< 0.001** |
| Female sex | 540 | | (57.7) | 438 | (58.2) | | 102 | | (55.4) | | 0.489 |
| *Comorbidities* |  | |  |  |  | |  | |  | |  |
| High blood pressure | 294 | | (31.4) | 164 | (21.8) | | 130 | | (70.6) | | **< 0.001** |
| Dyslipidaemia | 91 | | (9.7) | 33 | (4.4) | | 58 | | (31.5.) | | **< 0.001** |
| Chronic kidney disease | 68 | | (7.3) | 28 | (3.7) | | 40 | | (21.7) | | **< 0.001** |
| Previous stroke | 55 | | (5.9) | 27 | (3.6) | | 28 | | (15.2) | | **< 0.001** |
| Ischemic heart disease | 34 | | (3.6) | 16 | (2.1) | | 18 | | (9.8) | | **< 0.001** |
| Congestive heart failure | 25 | | (2.7) | 13 | (1.7) | | 12 | | (6.5) | | **0.001** |
| Ischemic lower limb disease | 22 | | (2.4) | 8 | (1.1) | | 14 | | (7.6) | | **< 0.001** |
| Peptic ulcer disease | 22 | | (2.4) | 12 | (1.6) | | 10 | | (5.4) | | **0.005** |
| Charlson score (medians, Q_1_-Q_3_) | 1 | | (0 – 3) | 0 | (0 - 2) | | 5 | | (3 - 7) | | *****< 0.001** |
| *Treatments* |  | |  |  |  | |  | |  | |  |
| Antihypertensive drugs | 241 | | (25.8) | 127 | (16.9) | | 114 | | (62.0) | | **< 0.001** |
| Antiplatelet agents | 117 | | (12.5) | 54 | (7.2) | | 63 | | (34.2) | | **< 0.001** |
| Statins | 111 | | (11.9) | 42 | (5.6) | | 69 | | (37.5) | | **< 0.001** |
| Anticoagulant drugs | 40 | | (4.3) | 26 | (3.5) | | 14 | | (7.6) | | **0.013** |
| Lipic-lowering drugs | 4 | | (0.5) | 4 | (0.5) | | 0 | | (0.0) | | 1.000 |
| *Symptoms (within 7 days)* |  | |  |  |  | |  | |  | |  |
| Fever | 776 | | (82.9) | 611 | (81.3) | | 165 | | (89.7) | | **0.001** |
| Fatigue | 624 | | (66.7) | 487 | (64.8) | | 137 | | (74.5) | | **0.012** |
| Loss of appetite | 436 | | (46.6) | 326 | (43.4) | | 110 | | (59.8) | | **< 0.001** |
| Myalgia | 518 | | (55.3) | 430 | (57.2) | | 88 | | (47.8) | | **0.022** |
| Headache | 471 | | (50.3) | 389 | (51.7) | | 82 | | (44.6) | | 0.082 |
| Arthralgia | 394 | | (42.1) | 323 | (43.0) | | 71 | | (38.6) | | 0.282 |
| Backache | 172 | | (18.4) | 138 | (18.4) | | 34 | | (18.5) | | 0.968 |
| Retro-orbital pain | 177 | | (18.9) | 147 | (19.6) | | 30 | | (16.3) | | 0.314 |
| *Vital constants* (medians, Q_1_-Q_3_) |  | |  |  |  | |  | |  | |  |
| Temperature (°C) | 38.1 | | (37.2 - 38.9) | 38.1 | (37.1 - 38.9) | | 38.4 | | (37.4 - 39.1) | | **0.009** |
| Heart rate (ppm) | 90 | | (77 - 103) | 89 | (76 - 102) | | 96 | | (83 - 107) | | **0.021** |
| Systolic blood pressure (mmHg) | 123 | | (110 - 139) | 121 | (109 - 136) | | 132 | | (116- 150) | | **< 0.001** |
| Diastolic blood pressure (mmHg) | 72 | | (63 - 81) | 72 | (62 - 81) | | 72 | | (64 - 81) | | 0.989 |
| Mean blood pressure (mmHg) | 90 | | (80 - 100) | 89 | (79 - 98) | | 93 | | (83 - 104) | | **0.016** |
| *Clinical features at presentation* |  | |  |  |  |  | |  | | |  |
| Days from symptom onset |  | |  |  |  |  | |  | | |  |
| ≤ 3 | 602 | | (68.7) | 468 | (66.9) | 134 | | (75.7) | | | **0.025** |
| > 3 | 274 | | (31.3) | 231 | (33.1) | 43 | | (24.3) | | |  |
| Indicators of probable dengue |  | |  |  |  |  | |  | | |  |
| Body aches | 664 | | (70.9) | 540 | (71.8) | 124 | | (67.4) | | | 0.237 |
| Nausea/Vomiting | 381 | | (40.7) | 312 | (41.5) | 69 | | (37.5) | | | 0.323 |
| Leukopenia (< 1,5 G/l) | 337 | | (36.0) | 286 | (38.0) | 51 | | (27.7) | | | 0.009 |
| Thrombocytopenia (< 100 G/l) | 162 | | (21.7) | 124 | (21.0) | 38 | | (24.5) | | | 0.347 |
| Skin rash | 148 | | (15.8) | 135 | (17.9) | 13 | | (7.1) | | | **< 0.001** |
| Purpura | 67 | | (7.2) | 57 | (7.6) | 10 | | (5.4) | | | 0.312 |
| Warning signs |  | |  |  |  |  | |  | | |  |
| 0 | 606 | | (64.7) | 490 | (65.2) | 116 | | (63.0) | | | 0.861 |
| 1 | 251 | | (26.8) | 199 | (26.5) | 52 | | (28.3) | | |  |
| > 1 | 79 | | (8.4) | 63 | (8.4) | 16 | | (8.7) | | |  |
| Abdominal pain/tenderness | 244 | | (26.1) | 189 | (25.1) | 55 | | (29.9) | | | 0.188 |
| Mucosal bleed | 99 | | (10.6) | 84 | (11.2) | 15 | | (8.2) | | | 0.233 |
| Persistent vomiting | 51 | | (5.5) | 40 | (5.3) | 11 | | (6.0) | | | 0.724 |
| Clinical fluid accumulation, n=749 | 42 | | (5.6) | 38 | (5.1) | 12 | | (7.7) | | | 0.195 |
| Lethargia/restlessness | 15 | | (1.6) | 14 | (1.9) | 1 | | (0.5) | | | 0.233 |
| Number of dengue indicators *^†^* |  | |  |  |  |  | |  | | |  |
| None | 117 | | (12.5) | 94 | (12.5) | 23 | | (12.5) | | | 0.725 |
| One | 189 | | (22.3) | 148 | (19.7) | 41 | | (22.3) | | |  |
| Two or more | 630 | | (65.2) | 510 | (67.8) | 120 | | (65.2) | | |  |
| Other manifestations |  | |  |  |  |  | |  | | |  |
| Dehydration | 306 | | (32.7) | 237 | (31.5) | 69 | | (37.5) | | | 0.121 |
| Muco-cutaneous signs | 187 | | (20.0) | 169 | (22.5) | 18 | | (9.8) | | | **< 0.001** |
| Cardiorespiratory signs | 170 | | (18.2) | 125 | (16.6) | 45 | | (24.5) | | | **0.013** |
| Non severe bleeding | 141 | | (15.1) | 120 | (16.0) | 21 | | (11.4) | | | 0.122 |
| Altered mental status | 80 | | (8.6) | 44 | (5.9) | 36 | | (19.6) | | | **< 0.001** |
| Itching | 66 | | (7.1) | 59 | (7.9) | 7 | | (3.8) | | | 0.055 |
| Cough | 60 | | (6.4) | 43 | (5.7) | 17 | | (9.2) | | | 0.080 |
| *Biology at presentation* (medians, Q_1_-Q_3_) | | |  |  |  | |  | |  | |  |
| Active thromboplastin time ratio | 1.1 | | (1.0 - 1.3) | 1.1 | (1.0 - 1.3) | | 1.2 | | (1.0 - 1.2) | | 0.592 |
| Alkaline phosphatase (IU/l) | 64 | | (52 - 82) | 63 | (52 - 81) | | 73 | | (55 - 91) | | 0.125 |
| ALT (IU/l) | 26 | | (16 - 49) | 26 | (15 - 49) | | 40 | | (20 - 88) | | 0.310 |
| AST (IU/l) | 37 | | (24 - 72) | 36 | (24 - 71) | | 42 | | (24 - 79) | | 0.257 |
| C reactive protein (mg/l) | 10.8 | | (4.7 - 23.8) | 8.6 | (3.9 - 20.7) | | 18.3 | | (8.7 - 31.7) | | **< 0.001** |
| Calcium (mmol/l) | 2.3 | | (2.2 – 2.3) | 2.3 | (2.2 – 2.3) | | 2.3 | | (2.2 – 2.3) | | 0.129 |
| Chlorine (mmol/l) | 99 | | (96 - 101) | 100 | (97 - 102) | | 97 | | (94 - 100) | | **< 0.001** |
| CPK (IU/l) | 127 | | (80 - 230) | 126 | (80 - 219) | | 140 | | (76 - 275) | | 0.397 |
| Creatinine (µmol/l) | 85 | | (69 - 106) | 81 | (67 - 99) | | 104 | | (78 - 135) | | *****< 0.001** |
| Fibrinogen (g/l) | 3.6 | | (3.1 - 4.2) | 3.5 | (3.1 - 4.1) | | 4.0 | | (3.5 - 4.5) | | **< 0.001** |
| Haematocrit (%) | 40.0 | | (36.4 - 43.4) | 40.5 | (37.1 - 43.8) | 37.9 | | (34.8 - 41.7) | | | **< 0.001** |
| Haemoglobin (g/dl) | 13.5 | | (12.3 - 14.9) | 13.7 | (12.5 - 15.1) | 12.7 | | (11.6 - 14.2) | | | **< 0.001** |
| International normalized ratio | 1.0 | | (1.0 - 1.1) | 1.0 | (1.0 - 1.1) | | 1.1 | | (1.0 - 1.1) | | **0.045** |
| Leucocytes (G/l) | 4.2 | | (2.9 - 6.0) | 4.0 | (2.8 - 5.6) | 5.0 | | (3.5 - 7.5) | | | **< 0.001** |
| Lipase (IU/l) | 33.9 | | (23.3 - 50.5) | 32.6 | (23.2 - 47.9) | | 38.5 | | (24.0 - 58.3) | | ****0.028** |
| Lymphocytes (G/l) | 0.6 | | (0.4 - 1.0) | 0.7 | (0.5 - 1.0) | 0.6 | | (0.4 - 0.9) | | | 0.247 |
| Monocyte to platelet ratio | 3.1 | | (2.0 - 5.0) | 2.9 | (1.8 - 4.6) | 3.8 | | (2.5 - 6.7) | | | ***0.001** |
| Monocytes (G/l) | 0.5 | | (0.3 - 0.7) | 0.5 | (0.3 - 0.7) | 0.6 | | (0.4 - 0.8) | | | **< 0.001** |
| Neutrophil to lymphocyte ratio | 4.2 | | (2.2 - 8.7) | 3.8 | (2.0 - 7.7) | | 5.5 | | (2.8 - 10.7) | | ***0.008** |
| Neutrophil to lymphocyte*platelet ratio | 3.1 | | (1.6 - 5.9) | 2.9 | (1.4 - 5.6) | | 4.3 | | (2.4 – 7.7) | | **< 0.001** |
| Neutrophil to platelet ratio | 19.4 | | (11.8 - 30.3) | 18.2 | (11.0 - 28.2) | | 25.8 | | (16.5 - 38.7) | | **< 0.001** |
| Neutrophils (G/l) | 2.8 | | (1.7 - 4.5) | 2.6 | (1.6 - 4.2) | 3.4 | | (2.1 - 5.4) | | | ***0.002** |
| Phosphate (mmol/l) | 1.0 | | (0.8 - 1.2) | 1.0 | (0.8 – 1.1) | | 1.0 | | (0.8 - 1.2) | | 0.390 |
| Platelet to lymphocyte ratio | 240 | | (140 - 367) | 235 | (144 - 350) | | 255 | | (128 - 397) | | 0.228 |
| Platelets (G/l) | 154 | | (107 - 202) | 157 | (109 - 204) | 152 | | (101 - 195) | | | 0.397 |
| Potassium (mmol/l) | 3.9 | | (3.6 - 4.2) | 3.8 | (3.6 – 4.1) | | 4.1 | | (3.7 – 4.5) | | ****< 0.001** |
| Prothrombin time (%) | 93 | | (82 - 101) | 93 | (83 - 101) | | 90 | | (81 - 101) | | 0.089 |
| Sodium (mmol/l) | 137 | | (134 - 139) | 137 | (135 - 139) | | 136 | | (133 - 138) | | **< 0.001** |
| Total bilirubin (mg/l) | 7.1 | | (4.9 - 10.7) | 7.1 | (4.8 - 10.2) | | 7.2 | | (4.9 - 12.3) | | 0.914 |
| Troponin (ng/l) | 0.1 | | (0.1 - 0.3) | 0.1 | (0.1 - 0.3) | | 0.2 | | (0.1 - 0.6) | | **0.019** |
| Urea (mmol/l) | 4.6 | | (3.3 – 6.5) | 4.2 | (3.0 - 5.8) | | 6.5 | | (4.8 - 9.6) | | *****< 0.001** |
| Urea to creatinine ratio | 52 | | (41 - 65) | 50 | (40 - 63) | | 60 | | (49 - 76) | | **< 0.001** |
| *Biology on hospital stay* (medians, Q_1_-Q_3_) | | |  |  |  | |  | |  | |  |
| Haemoconcentration | 42 | | (4.9) | 30 | (4.4) | | 12 | | (6.8) | | 0.195 |
| Platelet nadir (G/l) | 131 | | (70 - 180) | 136 | (84 - 184) | | 108 | | (42 – 60) | | **< 0.001** |
| Thrombocytopenia (< 100 G/l) | 303 | | (35.6) | 218 | (32.3) | | 85 | | (48.3) | | **< 0.001** |
| Albumin nadir (g/l), n=170 | 37.8 | | (34.4 - 40.9) | 38.0 | (34.5 - 41.0) | | 36.8 | | (34.0 - 39.7) | | 0.429 |
| HbA1c (%), n=76 | 7.2 | | (6.4 - 8.6) | 6.1 | (5.5 - 6.5) | | 7.8 | | (6.6 - 9.0) | | *****< 0.001** |
| Cholesterol total (g/l), n=111 | 7.2 | | (6.4 - 8.6) | 3.6 | (3.0 - 4.4) | | 3.1 | | (2.6 - 3.5) | | **< 0.001** |
| Triglycerides (g/l), n=111 | 1.9 | | (1.1 - 2.6) | 1.9 | (1.1 - 2.5) | | 1.9 | | (1.3 - 2.8) | | **0.946** |
| Peak troponin (ng/l), n=169 | 0.2 | | (0.1 - 0.3) | 0.1 | (0.0 - 0.3) | | 0.2 | | (0.1 - 0.8) | | **0.001** |
| Peak C Reactive protein (mg/l) | 12.6 | | (5.2 - 8.5) | 10.3 | (4.1 - 23.9) | | 21.2 | | (10.9 - 42.3) | | **< 0.001** |
| *Hospital outcomes* |  | |  |  |  | |  | |  | |  |
| Severe dengue (WHO 2009) | 185 | | (19.8) | 125 | (16.6) | | 60 | | (32.6) | | **< 0.001** |
| Co-infection | 103 | | (11.0) | 70 | (9.3) | | 33 | | (17.9) | | **0.001** |
| Hospitalization | 403 | | (43.1) | 287 | (38.2) | | 116 | | (63.0) | | **< 0.001** |
| LOS (days ; medians, Q_1_-Q_3_) | 4 | | (3 - 7) | 4 | (2 - 6) | | 6 | | (4 - 9) | | **< 0.001** |
| Hospitalization in the ICU | 64 | | (6.8) | 42 | (5.6) | | 22 | | (12.0) | | **0.002** |
| Death or need for critical care | 16 | | (1.7) | 7 | (0.9) | | 9 | | (5.0) | | **0.001** |
| *†This variable sums the numbers of probable dengue indicators and warning signs. Data are numbers and column percentages, or medians and interquartile ranges (Q1-Q3) when specified. Percentages are compared using chi2 or Fisher’s Exact test, as appropriate. Medians are compared using a non-parametric Brown-Mood k-sample test and when it was significant, further with median and quartile regressions. * p< 0.05, ** p<0.01 and *** p<0.001 for the less significant of both quantile regressions (no * means that at least one quantile regression is not significant)*. | | | | | | | | | | | |

**S2 Table. Characteristics of virologically confirmed cases of dengue among diabetic patients with or without micro- or macrovascular complications, Reunion island, January to June 2019.**

| **Variable** | **Non-diabetic**  **patients (NDPs) ^c^** | | **Diabetic patients (DPs)** | | | | | **^a versus b^** |
| --- | --- | --- | --- | --- | --- | --- | --- | --- |
|  |  |  | **Without diabetes complications ^b^** | | | **With diabetes complications ^b^** | |  |
|  | n=752 | (%) | | n=130 | (%) | n=54 |  | *p value* |
| *Socio-demographic* |  |  | |  |  |  |  |  |
| Age (years ; medians, Q_1_-Q_3_) | 45.6 | (29.6 - 63.1) | | ***68.5 | (55.9 - 76.0) | ***75.7 | (64.5 - 80.8) | **0.035** |
| Female sex | 438 | (58.2) | | 70 | (53.9) | 32 | (59.3) | 0.501 |
| *Comorbidities* |  |  | |  |  |  |  |  |
| High blood pressure | 164 | (21.8) | | ***79 | (60.8) | ***51 | (94.4) | **< 0.001** |
| Dyslipidaemia | 33 | (4.4) | | ***32 | (24.6) | ***26 | (48.2) | **0.002** |
| Chronic kidney disease | 28 | (3.7) | | *10 | (7.7) | ***30 | (55.6) | **< 0.001** |
| Previous stroke | 27 | (3.6) | | ***17 | (13.1) | ***11 | (20.4) | 0.210 |
| Ischemic heart disease | 16 | (2.1) | | ***11 | (8.5) | ***7 | (13.0) | 0.349 |
| Congestive heart failure | 13 | (1.7) | | 2 | (1.5) | ***10 | (18.5) | **< 0.001** |
| Peptic ulcer disease | 12 | (1.6) | | **8 | (6.2) | 2 | (3.7) | 0.726 |
| Ischemic lower limb disease | 8 | (1.1) | | 4 | (3.1) | ***10 | (18.5) | **0.001** |
| Charlson score (medians, Q_1_-Q_3_) | 0 | (0 - 2) | | ***4 | (2 - 5) | ***8 | (7 - 9) | **< 0.001** |
| *Treatments* |  |  | |  |  |  |  |  |
| Antihypertensive drugs | 127 | (16.9) | | ***73 | (56.2) | ***41 | (75.9) | 0.012 |
| Antiplatelet agents | 54 | (7.2) | | ***33 | (25.4) | ***30 | (55.6) | **< 0.001** |
| Statins | 42 | (5.6) | | ***38 | (29.2) | ***31 | (57.4) | **< 0.001** |
| Anticoagulant drugs | 26 | (3.5) | | 7 | (5.4) | **7 | (13.0) | 0.122 |
| Metformin | - | - | | 60 | (46.1) | 21 | (24.1) | **0.005** |
| Insulin | - | - | | 39 | (30.0) | 38 | (70.4) | **< 0.001** |
| *Symptoms (within 7 days)* |  |  | |  |  |  |  |  |
| Fever | 611 | (81.3) | | *110 | (88.5) | *50 | (92.6) | 0.402 |
| Fatigue | 487 | (64.8) | | 94 | (72.3) | *43 | (79.6) | 0.300 |
| Myalgia | 430 | (57.2) | | 64 | (49.2) | 24 | (44.4) | 0.554 |
| Headache | 389 | (51.7) | | 58 | (44.6) | 24 | (44.4) | 0.983 |
| Loss of appetite | 326 | (43.4) | | **77 | (59.2) | *33 | (61.1) | 0.813 |
| Arthralgia | 323 | (43.0) | | 52 | (41.9) | 19 | (31.7) | 0.180 |
| Retro-orbital pain | 147 | (19.6) | | 18 | (13.8) | 12 | (22.2) | 0.161 |
| Backache | 138 | (18.4) | | 23 | (17.7) | 11 | (20.4) | 0.677 |
| *Vital constants* (medians, Q_1_-Q_3_) |  |  | |  |  |  |  |  |
| Temperature (°C) | 38.1 | (37.1 - 38.9) | | *38.4 | (37.4 - 39.1) | 38.4 | (37.4 - 39.0) | 0.823 |
| Heart rate (ppm) | 89 | (76 - 102) | | *96 | (85 - 107) | 90 | (74 - 105) | 0.235 |
| Systolic blood pressure (mmHg) | 121 | (109 - 136) | | **133 | (116 - 147) | **131 | (112 - 158) | 0.961 |
| Diastolic blood pressure (mmHg) | 72 | (62 - 81) | | 74 | (66 - 82) | 70 | (58 - 80) | 0.155 |
| Mean blood pressure (mmHg) | 89 | (79 - 98) | | **93 | (85 - 102) | 90 | (79 - 105) | 0.928 |
| *Clinical features at presentation* |  |  | |  |  |  |  |  |
| Days from symptom onset |  |  | |  |  |  |  |  |
| ≤ 3 | 134 | (75.7) | | 94 | (75.6) | 40 | (76.8) | 0.808 |
| > 3 | 43 | (24.3) | | 31 | (24.8) | 12 | (23.1) |  |
| Indicators of probable dengue |  |  | |  |  |  |  |  |
| Body aches | 540 | (71.8) | | 91 | (70.0) | 33 | (61.1) | 0.242 |
| Nausea/Vomiting | 312 | (41.5) | | 49 | (37.7) | 20 | (37.0) | 0.933 |
| Leukopenia (< 1,5 G/l) | 286 | (38.0) | | *37 | (28.5) | 14 | (25.9) | 0.726 |
| Thrombocytopenia (< 100 G/l) | 162 | (21.7) | | 31 | (29.2) | 7 | (14.3) | **0.044** |
| Skin rash | 135 | (17.9) | | **11 | (8.5) | **2 | (3.7) | 0.351 |
| Purpura | 57 | (7.6) | | 4 | (3.1) | 6 | (11.1) | 0.066 |
| Warning signs |  |  | |  |  |  |  |  |
| 0 | 490 | (65.2) | | 81 | (62.3) | 35 | (64.8) | 0.080 |
| 1 | 199 | (26.5) | | 34 | (26.5) | 18 | (33.3) |  |
| > 1 | 63 | (8.4) | | 15 | (11.5) | 1 | (1.9) |  |
| Abdominal pain/tenderness | 189 | (25.1) | | 39 | (30.0) | 16 | (29.6) | 0.960 |
| Mucosal bleed | 84 | (11.2) | | 14 | (10.8) | *1 | (1.8) | 0.071 |
| Persistent vomiting | 40 | (5.3) | | 9 | (6.9) | 2 | (3.7) | 0.513 |
| Clinical fluid accumulation, n=700 | 30 | (5.1) | | 6 | (5.7) | *6 | (12.2) | 0.197 |
| Lethargia/restlessness | 14 | (1.9) | | 1 | (0.8) | 0 | (0.0) | 1.000 |
| Number of dengue indicators ^†^ |  |  | |  |  |  |  |  |
| None | 94 | (12.5) | | 16 | (12.3) | 7 | (13.0) | 0.731 |
| One | 148 | (19.7) | | 31 | (23.8) | 10 | (18.5) |  |
| Two or more | 510 | (67.8) | | 83 | (63.9) | 37 | (68.5) |  |
| Other manifestations |  |  | |  |  |  |  |  |
| Dehydration | 237 | (31.5) | | 43 | (33.1) | *26 | (48.2) | 0.054 |
| Muco-cutaneous signs | 169 | (22.5) | | **13 | (10.0) | *5 | (9.3) | 0.878 |
| Cardiorespiratory signs | 125 | (16.6) | | 29 | (22.3) | *16 | (29.6) | 0.293 |
| Non severe bleeding | 120 | (16.0) | | 18 | (13.9) | *3 | (5.6) | 0.131 |
| Itching | 59 | (7.9) | | 5 | (3.8) | 2 | (3.7) | 1.000 |
| Altered mental status | 44 | (5.9) | | ***25 | (19.2) | ***11 | (20.4) | 0.859 |
| Cough | 43 | (5.7) | | 13 | (10.0) | 4 | (7.4) | 0.781 |
| *Biology at presentation* (medians, Q_1_-Q_3_) | |  | |  |  |  |  |  |
| Active thromboplastin time ratio | 1.1 | (1.0 - 1.3) | | 1.1 | (1.0 - 1.2) | 1.2 | (1.0 - 1.2) | 0.923 |
| Alkaline phosphatase (IU/l) | 63 | (52 - 81) | | 72 | (56 - 85) | 76 | (55 - 114) | 0.362 |
| ALT (IU/l) | 26 | (15 - 49) | | 29 | (20 - 54) | 19 | (14 - 50) | **0.029** |
| AST (IU/l) | 36 | (24 - 71) | | 43 | (27 - 84) | 34 | (22 - 75) | 0.159 |
| C reactive protein (mg/l) | 8.6 | (3.9 - 20.7) | | ***16.9 | (8.6 - 31.0) | ***18.8 | (10.6 - 36.2) | 0.600 |
| Calcium (mmol/l) | 2.3 | (2.2 – 2.3) | | 2.3 | (2.2 - 2.4) | 2.3 | (2.1 – 2.3) | 0.351 |
| Chlorine (mmol/l) | 100 | (97 - 102) | | ***97 | (93 - 100) | *97 | (95 - 100) | 0.476 |
| CPK (IU/l) | 126 | (80 - 219) | | 130 | (78 - 235) | 163 | (74 - 298) | 0.479 |
| Creatinine (µmol/l) | 81 | (67 - 99) | | ***94 | (74 - 121) | ***131 | (101 - 172) | **< 0.001** |
| Fibrinogen (g/l) | 3.5 | (3.1 - 4.1) | | **4.0 | (3.4 - 4.4) | ***4.2 | (3.5 - 4.8) | 0.141 |
| Haematocrit (%) | 40.5 | (37.1 - 43.8) | | *39.1 | (35.6 - 42.5) | ***36.6 | (32.5 - 37.9) | **< 0.001** |
| Haemoglobin (g/dl) | 13.7 | (12.5 - 15.1) | | *13.2 | (11.8 - 14.5) | ***12.0 | (10.7 - 12.7) | **< 0.001** |
| International normalized ratio | 1.0 | (1.0 - 1.1) | | 1.1 | (1.0 - 1.1) | 1.1 | (1.0 - 1.2) | 0.446 |
| Leucocytes (G/l) | 4.0 | (2.8 - 5.6) | | **4.8 | (3.3 - 6.8) | **5.4 | (3.8 - 8.3) | 0.610 |
| Lipase (IU/l) | 32.6 | (23.2 - 47.9) | | *39.0 | (26.3 - 58.3) | 36.0 | (20.8 - 57.0) | 0.471 |
| Lymphocytes (G/l) | 0.7 | (0.5 - 1.0) | | 0.6 | (0.4 - 0.9) | 0.6 | (0.4 - 1.1) | 0.719 |
| Monocytes (G/l) | 0.5 | (0.3 - 0.7) | | **0.6 | (0.3 - 0.8) | ***0.5 | (0.3 - 0.9) | 0.559 |
| Monocyte to platelet ratio | 2.9 | (1.8 - 4.6) | | ***4.0 | (2.6 - 6.7) | 3.8 | (2.4 - 6.3) | 0.771 |
| Neutrophils (G/l) | 2.6 | (1.6 - 4.2) | | *3.3 | (2.0 - 4.8) | ***4.3 | (2.5 - 6.1) | 0.383 |
| Neutrophil to lymphocyte ratio | 3.8 | (2.0 - 7.7) | | *5.0 | (2.6 - 10.0) | **5.7 | (3.2 - 12.1) | 0.861 |
| Neutrophil to platelet ratio | 18.2 | (11.0 - 28.2) | | ***27.2 | (17.0 - 43.9) | **23.3 | (16.0 - 33.6) | 0.326 |
| Neutrophil to lymphocyte*platelet ratio | 2.9 | (1.4 - 5.6) | | 4.4 | (2.3 - 8.7) | 3.9 | (2.5 – 7.3) | 0.383 |
| Phosphate (mmol/l) | 1.0 | (0.8 – 1.1) | | 1.0 | (0.8 - 1.2) | 1.1 | (0.9 - 1.3) | 0.221 |
| Platelet to lymphocyte ratio | 235 | (144 - 350) | | 255 | (116 - 408) | 258 | (141 - 388) | 0.861 |
| Platelets (G/l) | 157 | (109 - 204) | | 146 | (87 - 192) | 162 | (131 - 206) | 0.230 |
| Potassium (mmol/l) | 3.8 | (3.6 – 4.1) | | **4.1 | (3.6 - 4.4) | ***4.2 | (3.8 - 4.7) | 0.630 |
| Prothrombin time (%) | 93 | (83 - 101) | | 90 | (81 - 103) | 90 | (79 - 99) | 0.939 |
| Sodium (mmol/l) | 137 | (135 - 139) | | **135 | (132 - 138) | *136 | (134 - 138) | 0.245 |
| Total bilirubin (mg/l) | 7.1 | (4.8 - 10.2) | | 8.1 | (5.2 - 13.4) | 6.8 | (4.3 - 10.9) | 0.696 |
| Troponin (ng/l) | 0.1 | (0.1 - 0.3) | | 0.1 | (0.1 - 0.2) | ***0.3 | (0.1 - 0.9) | 0.141 |
| Urea (mmol/l) | 4.2 | (3.0 - 5.8) | | ***5.8 | (4.5 - 8.0) | ***7.7 | (5.6 - 12.4) | 0.057 |
| Urea to creatinine ratio | 50 | (40 - 63) | | ***60 | (49 - 77) | *61 | (48 - 74) | 0.608 |
| *Biology on hospital stay* (medians, Q_1_-Q_3_) | |  | |  |  |  |  |  |
| Haemoconcentration | 30 | (4.4) | | 6 | (4.9) | *6 | (11.3) | 0.188 |
| Platelet nadir (G/l) | 136 | (84 - 184) | | *108 | (36 - 163) | *96 | (55 - 151) | 0.511 |
| Thrombocytopenia (< 100 G/l) | 85 | (48.3) | | 58 | (47.2) | 27 | (50.9) | 0.644 |
| Albumin nadir (g/l), n=170 | 38.0 | (34.5 - 41.0) | | 37.4 | (34.7 - 39.7) | 36.2 | (32.4 - 40.0) | 0.439 |
| HbA1c (%), n=76 | 6.1 | (5.5 - 6.5) | | **8.0 | (6.5 - 9.7) | ***7.5 | (7.0 - 8.2) | 0.917 |
| Cholesterol total (g/l), n=111 | 3.6 | (3.0 - 4.4) | | 3.2 | (2.7 - 3.8) | *2.8 | (2.6 - 3.2) | 0.851 |
| Triglycerides (g/l), n=111 | 1.9 | (1.1 - 2.5) | | 2.2 | (1.6 - 2.8) | 1.5 | (1.1 - 2.4) | 0.394 |
| Peak troponin (ng/l), n=169 | 0.1 | (0.0 - 0.3) | | 0.2 | (0.1 - 0.3) | ***0.3 | (0.1 - 1.1) | 0.137 |
| Peak C Reactive protein (mg/l) | 10.3 | (4.1 - 23.9) | | ***20.5 | (10.3 - 37.5) | ***32.3 | (14.7 - 60.6) | **< 0.001** |
| *Hospital outcomes* |  |  | |  |  |  |  |  |
| Severe dengue (WHO 2009) | 125 | (16.6) | | **34 | (26.1) | ***26 | (48.2) | **0.004** |
| Co-infection | 70 | (9.3) | | *21 | (16.1) | **12 | (22.2) | 0.329 |
| Hospitalization | 287 | (38.2) | | ***73 | (56.1) | ***43 | (79.6) | **0.003** |
| LOS (days; medians, Q_1_-Q_3_) | 4 | (2 - 6) | | **5 | (3 - 9) | ***6 | (4 - 9) | 0.562 |
| Hospitalization in the ICU | 42 | (5.6) | | **16 | (12.3) | 6 | (11.1) | 0.820 |
| Death or need for critical care | 7 | (0.9) | | 4 | (3.1) | **5 | (9.3) | 0.128 |
| *^†^This variable sums the numbers of probable dengue indicators and warning signs. ALT: alanine aminotransferase. AST: aspartate aminotransferase. CPK: creatine phosphor kinase. HbA1c : glycosylated haemoglobin. LOS: length of hospital stay. ICU: intensive care unit. Data are numbers and column percentages, or medians and interquartile ranges (Q1-Q3) when specified. Percentages are compared using chi2 or Fisher’s Exact test, as appropriate. Medians are compared using a non-parametric Brown-Mood k-sample test and when it was significant, further with median and quartile regressions. Exact p values are given for column c versus b. P values are only expressed as * for column b vs a and column c vs a. * p< 0.05, ** p<0.01 and *** p<0.001 for the less significant of both quantile regressions (no * means that at least one quantile regression is not significant).* | | | | | | | | |

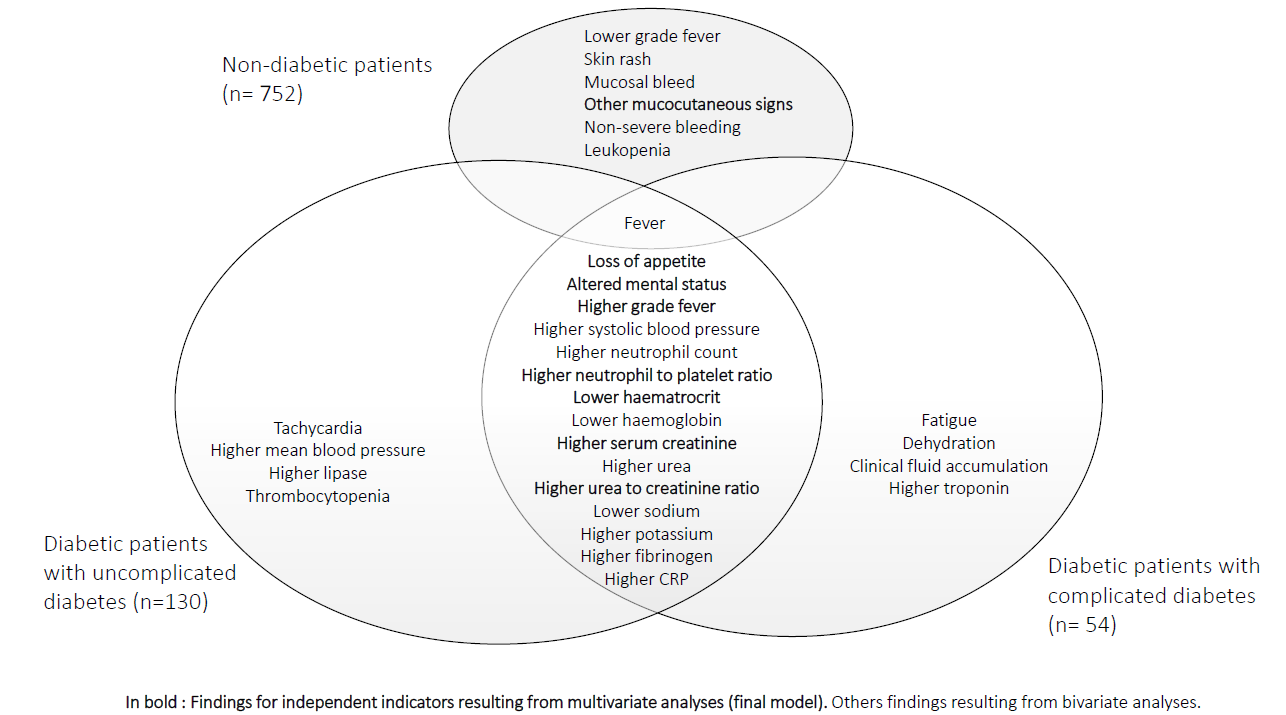

**S1 Figure. Indicators of dengue in the non-diabetic patients (NDPs), in the diabetic patients (DPs) with uncomplicated diabetes and with diabetes complications.**

| **S3 Table. Characteristics of virologically confirmed cases of dengue among diabetic patients with optimal and suboptimal glycaemic control, Reunion island, January to June 2019.** | | | | | | | | | | | |
| --- | --- | --- | --- | --- | --- | --- | --- | --- | --- | --- | --- |
| **Variable** | **Non-diabetic patients ^c^** | | | **HBA1c < 7% ^b^** | | | **HBA1c ≥ 7% ^a^** | | | **^a versus b^** | |
|  | | n=752 | (%) | n=17 | (%) | | n=42 | (%) | | | *p value* |
| *Socio-demographic* | |  |  |  |  | |  |  | | |  |
| Age (years ; medians, Q_1_-Q_3_) | | 45.6 | (29.6 - 63.1) | ***75.5 | (64.9 - 79.4) | | ***74.7 | (67.8 - 79.1) | | | 0.934 |
| Female sex | | 438 | (58.2) | 8 | (44.4) | | 22 | (52.4) | | | 0.711 |
| *Comorbidities* | |  | 5 |  |  | |  |  | | |  |
| Advanced/complicated diabetes | | 0 | (0.0) | 3 | (17.6) | | *20 | (47.6) | | | 0.042 |
| High blood pressure | | 164 | (21.8) | ***12 | (66.7) | | ***34 | (80.9) | | | 0.197 |
| Dyslipidaemia | | 33 | (4.4) | **5 | (27.8) | | ***15 | (35.7) | | | 0.643 |
| Chronic kidney disease | | 28 | (3.7) | 3 | (16.7) | | ***13 | (30.9) | | | 0.190 |
| Previous stroke | | 27 | (3.6) | 1 | (5.6) | | **7 | (16.7) | | | 0.417 |
| Ischemic heart disease | | 16 | (2.1) | 0 | (0.0) | | *4 | (9.5) | | | 0.314 |
| Congestive heart failure | | 13 | (1.7) | 0 | (0.0) | | *3 | (7.1) | | | 0.550 |
| Peptic ulcer disease | | 12 | (1.6) | 3 | (16.7) | | *4 | (9.5) | | | 0.399 |
| Ischemic lower limb disease | | 8 | (1.1) | 1 | (5.6) | | **3 | (7.1) | | | 1.000 |
| Charlson score (medians, Q_1_-Q_3_) | | 0 | (0 - 2) | ***5 | (4 - 6) | | ***6 | (4 - 8) | | | 0.091 |
| *Treatments* | |  |  |  |  | |  |  | | |  |
| Antihypertensive drugs | | 127 | (16.9) | **10 | (55.6) | | ***28 | (66.7) | | | 0.323 |
| Antiplatelet agents | | 54 | (7.2) | **7 | (38.9) | | ***18 | (42.9) | | | 0.592 |
| Statins | | 42 | (5.6) | ***7 | (38.9) | | ***20 | (47.6) | | | 0.653 |
| Anticoagulant drugs | | 26 | (3.5) | 3 | (16.7) | | 2 | (4.8) | | | 0.138 |
| Metformin | | - | - | 7 | (38.9) | | 15 | (35.7) | | | 0.694 |
| Insulin | | - | - | 6 | (33.3) | | 19 | (45.2) | | | 0.262 |
| *Symptoms (within 7 days)* | |  |  |  |  | |  |  | | |  |
| Fever | | 611 | (81.3) | 16 | (94.1) | | *40 | (95.2) | | | 1.000 |
| Fatigue | | 487 | (64.8) | *16 | (94.1) | | 37 | (88.1) | | | 0.662 |
| Myalgia | | 430 | (57.2) | 11 | (64.7) | | 21 | (50.0) | | | 0.304 |
| Headache | | 389 | (51.7) | 9 | (52.9) | | 16 | (38.1) | | | 0.296 |
| Loss of appetite | | 326 | (43.4) | 11 | (64.1) | | ***32 | (76.2) | | | 0.519 |
| Arthralgia | | 323 | (43.0) | 9 | (52.9) | | 17 | (40.5) | | | 0.382 |
| Retro-orbital pain | | 147 | (19.6) | 3 | (17.7) | | 5 | (11.9) | | | 0.678 |
| Backache | | 138 | (18.4) | 3 | (17.7) | | 7 | (16.7) | | | 1.000 |
| *Vital constants* (medians, Q_1_-Q_3_) | |  |  |  |  | |  |  | | |  |
| Temperature (°C) | | 38.1 | (37.1 - 38.9) | 38.6 | (37.7 - 39.2) | | 38.4 | (37.1 - 39.1) | | | 0.934 |
| Heart rate (ppm) | | 89 | (76 - 102) | 95 | (78 - 97) | | 91 | (76 - 104) | | | 0.934 |
| Systolic blood pressure (mmHg) | | 121 | (109 - 136) | 123 | (106 - 139) | | *129 | (112 - 145) | | | 0.934 |
| Diastolic blood pressure (mmHg) | | 72 | (62 - 81) | 69 | (64 - 81) | | 70 | (58 - 77) | | | 0.804 |
| Mean blood pressure (mmHg) | | 89 | (79 - 98) | 91 | (78 - 99) | | 89 | (79 - 99) | | | 0.934 |
| *Clinical features at presentation* | |  |  |  |  | |  |  | | |  |
| Days from symptom onset | |  |  |  |  | |  |  | | |  |
| ≤ 3 | | 468 | (66.9) | 16 | (94.1) | | 32 | (76.2) | | | 0.265 |
| > 3 | | 231 | (33.1) | 1 | (5.9) | | 10 | (23.8) | | |  |
| Indicators of probable dengue | |  |  |  |  | |  |  | | |  |
| Body aches | | 540 | (71.8) | 13 | (76.5) | | 29 | (69.0) | | | 0.753 |
| Nausea/Vomiting | | 312 | (41.5) | 6 | (35.3) | | 14 | (33.3) | | | 0.852 |
| Leukopenia (< 1,5 G/l) | | 286 | (38.0) | 8 | (47.1) | | 18 | (42.9) | | | 0.768 |
| Thrombocytopenia (< 100 G/l) | | 162 | (21.7) | 4 | (26.7) | | *15 | (37.5) | | | 0.537 |
| Skin rash | | 135 | (17.9) | 2 | (11.8) | | *2 | (4.8) | | | 0.571 |
| Purpura | | 57 | (7.6) | 1 | (5.9) | | 0 | (0.0) | | | 0.305 |
| Warning signs | |  |  |  |  | |  |  | | |  |
| 0 | | 490 | (65.2) | 12 | (70.6) | | 26 | (61.9) | | | 0.287 |
| 1 | | 199 | (26.5) | 3 | (17.7) | | 14 | (33.3) | | |  |
| > 1 | | 63 | (8.4) | 2 | (11.8) | | 2 | (4.8) | | |  |
| Abdominal pain/tenderness | | 189 | (25.1) | 4 | (23.5) | | 11 | (26.2) | | | 1.000 |
| Mucosal bleed | | 84 | (11.2) | 2 | (11.8) | | 5 | (11.9) | | | 1.000 |
| Persistent vomiting | | 40 | (5.3) | 1 | (5.9) | | 1 | (2.4) | | | 0.497 |
| Clinical fluid accumulation, n=634 | | 30 | (5.1) | 0 | (0.0) | | 4 | (10.3) | | | 1.000 |
| Lethargia/restlessness | | 14 | (1.9) | 0 | (0.8) | | 0 | (0.0) | | | N.A |
| Number of dengue indicators ^†^ | |  |  |  |  | |  |  | | |  |
| None | | 94 | (12.5) | 1 | (5.9) | | 5 | (11.9) | | | 0.884 |
| One | | 148 | (19.7) | 2 | (11.8) | | 5 | (11.9) | | |  |
| Two or more | | 510 | (67.8) | 14 | (82.4) | | 32 | (76.2) | | |  |
| Other manifestations | |  |  |  |  | |  |  | | |  |
| Dehydration | | 237 | (31.5) | 7 | (41.2) | | *20 | (47.6) | | | 0.653 |
| Muco-cutaneous signs | | 169 | (22.5) | 2 | (11.8) | | 5 | (11.9) | | | 1.000 |
| Cardiorespiratory signs | | 125 | (16.6) | 6 | (35.3) | | 11 | (26.2) | | | 0.535 |
| Non severe bleeding | | 120 | (16.0) | 2 | (11.8) | | 5 | (11.9) | | | 1.000 |
| Itching | | 59 | (7.9) | 1 | (5.9) | | 2 | (4.8) | | | 0.218 |
| Altered mental status | | 44 | (5.9) | 7 | (41.2) | | ***14 | *(33.3) | | | 0.347 |
| Cough | | 43 | (5.7) | 3 | (17.7) | | 4 | (9.5) | | | 0.393 |
| *Biology at presentation* (medians, Q_1_-Q_3_) | | |  |  |  | |  |  | | |  |
| Active thromboplastin time ratio | | 1.1 | (1.0 - 1.3) | 1.2 | (1.0 - 1.4) | | 1.2 | (1.0 - 1.2) | | | 0.486 |
| Alkaline phosphatase (IU/l) | | 63 | (52 - 81) | 68 | (57 - 91) | | 78 | (56 - 90) | | | 0.262 |
| ALT (IU/l) | | 26 | (15 - 49) | 26 | (20 - 56) | | 27 | (15 - 57) | | | 0.932 |
| AST (IU/l) | | 36 | (24 - 71) | 55 | (33 - 73) | | 43 | (23 - 83) | | | 0.689 |
| C reactive protein (mg/l) | | 8.6 | (3.9 - 20.7) | **20.8 | (10.6 - 51.0) | | ***19.0 | (12.8 - 33.9) | | | 0.931 |
| Calcium (mmol/l) | | 2.3 | (2.2 – 2.3) | 2.3 | (2.2 - 2.3) | | 2.3 | (2.1 – 2.3) | | | 0.140 |
| Chlorine (mmol/l) | | 100 | (97 - 102) | 99 | (96 - 1022) | | ***95 | (92 - 97) | | | 0.058 |
| CPK (IU/l) | | 126 | (80 - 219) | 207 | (108 - 342) | | 163 | (90 - 296) | | | 0.755 |
| Creatinine (µmol/l) | | 81 | (67 - 99) | *90 | (86 - 123) | | ***110 | (92 - 148) | | | 0.259 |
| Fibrinogen (g/l) | | 3.5 | (3.1 - 4.1) | *4.0 | (3.8 - 4.5) | | *4.0 | (3.5 - 4.5) | | | 0.941 |
| Haematocrit (%) | | 40.5 | (37.1 - 43.8) | 36.4 | (34.2 - 40.6) | | *36.8 | (33.6 - 42.2) | | | 0.934 |
| Haemoglobin (g/dl) | | 13.7 | (12.5 - 15.1) | 11.9 | (11.5 - 14.1) | | *12.5 | (11.1 - 14.2) | | | 0.601 |
| International normalized ratio | | 1.0 | (1.0 - 1.1) | *1.1 | (1.0 - 1.2) | | 1.0 | (1.0 - 1.1) | | | 0.602 |
| Leucocytes (G/l) | | 4.0 | (2.8 - 5.6) | 3.9 | (3.3 - 5.7) | | 4.6 | (3.0 - 7.3) | | | 0.720 |
| Lipase (IU/l) | | 32.6 | (23.2 - 47.9) | 48.3 | (22.3 - 108) | | 33.7 | (25.0 - 57.0) | | | 0.689 |
| Lymphocytes (G/l) | | 0.7 | (0.5 - 1.0) | 0.6 | (0.4 - 0.8) | | 0.6 | (0.5 - 1.0) | | | 0.795 |
| Monocytes (G/l) | | 0.5 | (0.3 - 0.7) | *0.6 | | (0.4 - 0.7) | 0.6 | | (0.4 - 0.9) | | 0.653 |
| Monocyte to platelet ratio | | 2.9 | (1.8 - 4.6) | 4.0 | | (2.4 - 4.5) | **5.3 | | (2.7 - 7.3) | | **0.019** |
| Neutrophils (G/l) | | 2.6 | (1.6 - 4.2) | 2.7 | (2.0 - 4.4) | | 2.6 | (1.8 - 5.1) | | | 0.858 |
| Neutrophil to lymphocyte ratio | | 3.8 | (2.0 - 7.7) | 4.2 | (3.0 - 8.3) | | 4.2 | (2.6 - 10.5) | | | 0.795 |
| Neutrophil to lymphocyte*platelet ratio | | 2.9 | (1.4 - 5.6) | 3.8 | (2.0 - 12.9) | | 4.0 | (2.5 - 7.5) | | | 0.931 |
| Neutrophil to platelet ratio | | 18.2 | (11.0 - 28.2) | 23.1 | (15.3 - 29.3) | | *25.6 | (14.5 - 45.2) | | | 0.934 |
| Phosphate (mmol/l) | | 1.0 | (0.8 – 1.1) | 1.1 | (0.9 - 1.2) | | 1.0 | (0.8 - 1.3) | | | 0.348 |
| Platelet to lymphocyte ratio | | 235 | (144 - 350) | 270 | (167 - 445) | | 215 | (119 - 324) | | | 0.522 |
| Platelets (G/l) | | 157 | (109 - 204) | 145 | (93 - 185) | | 142 | (77 - 166) | | | 0.486 |
| Potassium (mmol/l) | | 3.8 | (3.6 – 4.1) | 4.2 | (3.8 - 4.4) | | 4.3 | (3.8 - 4.7) | | | 0.516 |
| Prothrombin time (%) | | 93 | (83 - 101) | 84 | (75 - 91) | | 92 | (84 - 101) | | | 0.081 |
| Sodium (mmol/l) | | 137 | (135 - 139) | 136 | (135 - 139) | | ***134 | (131 - 136) | | | **0.019** |
| Total bilirubin (mg/l) | | 7.1 | (4.8 - 10.2) | 6.4 | (4.3 - 19.3) | | 6.5 | (4.7 - 10.9) | | | 0.911 |
| Troponin (ng/l) | | 0.1 | (0.1 - 0.3) | 0.1 | (0.1 - 0.5) | | 0.2 | (0.1 - 0.6) | | | 0.879 |
| Urea (mmol/l) | | 4.2 | (3.0 - 5.8) | 5.8 | (3.9 - 7.4) | | ***7.0 | (5.2 - 10.2) | | | 0.259 |
| Urea to creatinine ratio | | 50 | (40 - 63) | 51 | (46 - 67) | | **61 | (49 - 76) | | | 0.423 |
| *Biology on hospital stay* (medians, Q_1_-Q_3_) | | |  |  |  | |  |  | | |  |
| Haemoconcentration | | 30 | (4.4) | 1 | (5.9) | | 4 | (9.5) | | | 1.000 |
| Platelet nadir (G/l) | | 136 | (84 - 184) | *68 | (38 – 108) | | **62 | (23 - 136) | | | 0.934 |
| Thrombocytopenia (< 100 G/l) | | 42 | (71.2) | 12 | (70.6) | | 30 | (71.4) | | | 1.000 |
| Albumin nadir (g/l), n=170 | | 38.0 | (34.5 - 41.0) | 36.7 | (34.7 - 40.6) | | 37.2 | (33.0 - 39.3) | | | 0.808 |
| Cholesterol total (g/l), n=111 | | 3.6 | (3.0 - 4.4) | 3.3 | (2.8 - 3.6) | | **2.8 | (2.6 - 3.2) | | | 0.596 |
| Triglycerides (g/l), n=111 | | 1.9 | (1.1 - 2.5) | 1.9 | (1.2 - 2.9) | | 1.7 | (1.0 - 2.5) | | | 0.909 |
| Peak troponin (ng/l), n=169 | | 0.1 | (0.0 - 0.3) | 0.2 | (0.1 - 0.2) | | 0.2 | (0.1 - 0.7) | | | 0.875 |
| Peak C Reactive protein (mg/l) | | 10.3 | (4.1 - 23.9) | *37.9 | (12.3 - 58.4) | | ***24.7 | (14.7 - 40.0) | | | 0.934 |
| *Hospital outcomes* | |  |  |  |  | |  |  | | |  |
| Severe dengue (WHO 2009) | | 125 | (16.6) | ***11 | (64.7) | | ***20 | (47.6) | | | 0.234 |
| Co-infection | | 70 | (9.3) | 2 | (11.8) | | **11 | (26.2) | | | 0.313 |
| Hospitalization | | 287 | (38.2) | 17 | (100.0) | | 42 | (100.0) | | | 1.000 |
| LOS (days ; medians, Q_1_-Q_3_) | | 4 | (2 - 6) | 6 | **(5 - 10) | | 7 | (5 - 10) | | | 0.808 |
| Hospitalization in the ICU | | 42 | (5.6) | 4 | *(23.5) | | 4 | (9.5) | | | 0.211 |
| Death or need for critical care | | 7 | (0.9) | 1 | (5.6) | | 2 | (4.8) | | | 1.000 |
| *^†^This variable sums the numbers of probable dengue indicators and warning signs. ALT: alanine aminotransferase. AST: aspartate aminotransferase. CPK: creatine phosphor kinase. HbA1c : glycosylated haemoglobin. LOS: length of hospital stay. ICU: intensive care unit. Data are numbers and column percentages, or medians and interquartile ranges (Q1-Q3) when specified. Percentages are compared using chi2 or Fisher’s Exact test, as appropriate. Medians are compared using a non-parametric Brown-Mood k-sample test and when it was significant, further with median and quartile regressions. Exact p values are given for column c versus b. P values are only expressed as * for column b vs a and column c vs a. * p< 0.05, ** p<0.01 and *** p<0.001 for the less significant of both quantile regressions (no * means that at least one quantile regression is not significant).*  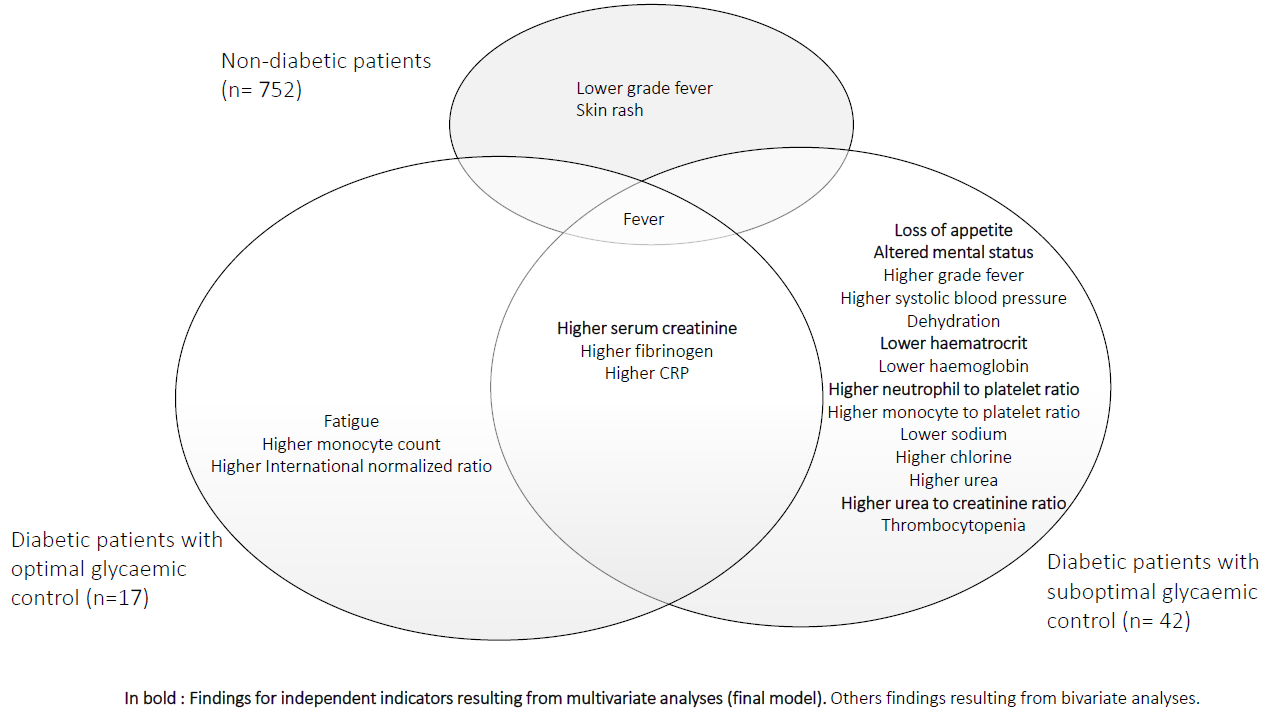 | | | | | | | | | | | |

**S2 Figure. Indicators of dengue in the non-diabetic patients (NDPs), in the diabetic patients (DPs) with optimal glycaemic control (< 7%) or with suboptimal glycaemic control (≥ 7%).**

| **S4 Table. Characteristics of virologically confirmed cases of non severe and severe dengue cases among diabetic patients, Reunion island, January to June 2019.** | | | | | | | | | |
| --- | --- | --- | --- | --- | --- | --- | --- | --- | --- |
| **Variable** | | **Total** | | **Non severe dengue (DF)** | | **Severe dengue (SD)** | | |  |
|  | n=184 | | (%) | n=124 | (%) | | n=60 | (%) | *p value* |
| *Socio-demographic* |  | |  |  |  | |  |  |  |
| Age (years ; medians, Q_1_-Q_3_) | 70.0 | | (59.4 - 77.7) | 70.0 | (58.6 - 77.3) | | 69.6 | (60.6 - 78.4) | 0.875 |
| Female sex | 102 | | (55.4) | 68 | (54.8) | | 34 | (56.7) | 0.815 |
| *Comorbidities* |  | |  |  |  | |  |  |  |
| High blood pressure | 130 | | (70.6) | 84 | (67.7) | | 46 | (76.7) | 0.213 |
| Dyslipidaemia | 58 | | (31.5) | 37 | (29.8) | | 21 | (35.0.) | 0.502 |
| Chronic kidney disease | 40 | | (21.7) | 21 | (16.9) | | 19 | (31.7) | **0.023** |
| Previous stroke | 28 | | (15.2) | 20 | (16.1) | | 8 | (13.3) | 0.621 |
| Ischemic heart disease | 18 | | (9.8) | 15 | (12.1) | | 3 | (5.0) | 0.186 |
| Congestive heart failure | 12 | | (6.5) | 6 | (4.8) | | 6 | (10.0) | 0.210 |
| Ischemic lower limb disease | 14 | | (7.6) | 8 | (6.4) | | 6 | (10.0) | 0.390 |
| Peptic ulcer disease | 10 | | (5.4) | 4 | (3.2) | | 6 | (10.0) | 0.081 |
| Charlson score (medians, Q_1_-Q_3_) | 5 | | (3 - 7) | 4 | (3 - 6) | | 6 | (3 - 8) | ***0.018** |
| *Treatments* |  | |  |  |  | |  |  |  |
| Antihypertensive drugs | 114 | | (62.0) | 77 | (62.1) | | 37 | (61.7) | 0.955 |
| Insulin | 77 | | (41.9) | 50 | (40.3) | | 27 | (45.0) | 0.547 |
| Metformin | 73 | | (39.7) | 52 | (41.9) | | 21 | (35.0) | 0.367 |
| Statins | 69 | | (37.5) | 46 | (37.1) | | 23 | (38.3) | 0.871 |
| Antiplatelet agents | 63 | | (34.2) | 40 | (32.3) | | 23 | (38.3) | 0.508 |
| Anticoagulant drugs | 14 | | (7.6) | 8 | (6.4) | | 6 | (10.0) | 0.390 |
| *Symptoms (within 7 days)* |  | |  |  |  | |  |  |  |
| Fever | 165 | | (89.7) | 110 | (88.7) | | 55 | (91.7) | 0.537 |
| Fatigue | 137 | | (74.5) | 88 | (71.0) | | 49 | (81.7) | 0.119 |
| Loss of appetite | 110 | | (59.8) | 70 | (56.5) | | 40 | (66.7) | 0.185 |
| Myalgia | 88 | | (47.8) | 64 | (51.6) | | 24 | (40.0) | 0.139 |
| Headache | 82 | | (44.6) | 61 | (49.2) | | 21 | (35.0) | 0.069 |
| Arthralgia | 71 | | (38.6) | 52 | (41.9) | | 19 | (31.7) | 0.180 |
| Backache | 34 | | (18.5) | 28 | (22.6) | | 6 | (10.0) | **0.039** |
| Retro-orbital pain | 30 | | (16.3) | 22 | (17.7) | | 8 | (13.3) | 0.448 |
| *Vital constants* (medians, Q_1_-Q_3_) |  | |  |  |  | |  |  |  |
| Temperature (°C) | 38.4 | | (37.4 - 39.1) | 38.4 | (37.4 - 39.2) | | 38.2 | (37.4 - 39.0) | 0.530 |
| Heart rate (ppm) | 96 | | (83 - 107) | 96 | (83 - 104) | | 96 | (83 - 112) | 0.588 |
| Systolic blood pressure (mmHg) | 132 | | (116 - 150) | 133 | (119 - 150) | | 131 | (110 - 152) | 0.957 |
| Diastolic blood pressure (mmHg) | 72 | | (64 - 81) | 71 | (64 - 81) | | 73 | (64 - 81) | 0.787 |
| Mean blood pressure (mmHg) | 93 | | (83 - 104) | 92 | (84 - 103) | 96 | | (79 - 104) | 0.121 |
| *Clinical features at presentation* |  | |  |  |  |  | |  |  |
| Days from symptom onset |  | |  |  |  |  | |  |  |
| ≤ 3 | 134 | | (75.7) | 80 | (72.3) | 44 | | (74.6) | 0.522 |
| > 3 | 43 | | (24.3) | 28 | (23.7) | 15 | | (25.4) |  |
| Indicators of probable dengue |  | |  |  |  |  | |  |  |
| Body aches | 124 | | (67.4) | 90 | (72.6) | 34 | | (56.7) | **0.031** |
| Nausea/Vomiting | 69 | | (37.5) | 45 | (36.3) | 24 | | (40.0) | 0.626 |
| Leukopenia (< 1,5 G/l) | 51 | | (27.7) | 33 | (26.6) | 18 | | (30.0) | 0.630 |
| Thrombocytopenia (< 100 G/l) | 38 | | (24.5) | 23 | (22.5) | 15 | | (28.3) | 0.430 |
| Skin rash | 13 | | (7.1) | 9 | (7.3) | 4 | | (6.7) | 1.000 |
| Purpura | 10 | | (5.4) | 6 | (4.8) | 4 | | (6.7) | 0.730 |
| Warning signs |  | |  |  |  |  | |  |  |
| 0 | 116 | | (63.0) | 83 | (66.9) | 33 | | (55.0) | 0.079 |
| 1 | 52 | | (28.3) | 34 | (27.4) | 18 | | (30.0) |  |
| > 1 | 16 | | (8.7) | 7 | (5.7) | 9 | | (15.0) |  |
| Abdominal pain/tenderness | 55 | | (29.1) | 34 | (27.4) | 21 | | (35.0) | 0.292 |
| Mucosal bleed | 15 | | (8.2) | 7 | (5.6) | 8 | | (13.3) | 0.088 |
| Clinical fluid accumulation (n=155) | 12 | | (7.7) | 3 | (2.9) | 9 | | (17.0) | **0.003** |
| Persistent vomiting | 11 | | (6.0) | 6 | (4.8) | 5 | | (8.3) | 0.342 |
| Lethargia/restlesness | 1 | | (0.5) | 1 | (0.8) | 0 | | (0.0) | 1.000 |
| Number of dengue indicators ^†^ |  | |  |  |  |  | |  |  |
| None | 23 | | (12.5) | 16 | (12.9) | 7 | | (11.6) | 0.391 |
| One | 41 | | (22.3) | 31 | (25.0) | 10 | | (16.7) |  |
| Two or more | 120 | | (65.2) | 77 | (62.1) | 43 | | (71.7) |  |
| Other manifestations |  | |  |  |  |  | |  |  |
| Dehydration | 69 | | (37.5) | 40 | (32.3) | 29 | | (48.3) | **0.035** |
| Cardiorespiratory signs | 45 | | (24.5) | 18 | (14.5) | 27 | | (45.0) | **< 0.001** |
| Altered mental status | 36 | | (19.6) | 18 | (14.5) | 18 | | (30.0) | **0.013** |
| Non severe bleeding | 21 | | (11.4) | 9 | (7.3) | 12 | | (20.0) | **0.011** |
| Muco-cutaneous signs | 18 | | (9.8) | 10 | (8.1) | 8 | | (13.3) | 0.259 |
| Cough | 17 | | (9.2) | 7 | (5.6) | 10 | | (16.7) | **0.016** |
| Itching | 7 | | (3.8) | 4 | (3.2) | 3 | | (5.0) | 0.684 |
| *Biology at presentation* (medians, Q_1_-Q_3_) | | |  |  |  |  | |  |  |
| Active thromboplastin time ratio | 1.1 | | (1.0 - 1.2) | 1.1 | (1.0 - 1.2) | 1.2 | | (1.0 - 1.2) | 0.625 |
| Alkaline phosphatase (IU/l) | 73 | | (55 - 91) | 62 | (54 - 87) | 80 | | (72 - 106) | **0.015** |
| ALT (IU/l) | 26 | | (16 - 52) | 23 | (14 - 43) | 37 | | (20 - 66) | 0.216 |
| AST (IU/l) | 42 | | (24 - 79) | 37 | (22 - 70) | 46 | | (31 - 103) | 0.070 |
| C reactive protein (mg/l) | 18.3 | | (8.7 - 31.7) | 16.3 | (8.0 - 29.5) | 21.2 | | (10.4 - 42.9) | 0.301 |
| Calcium (mmol/l) | 2.3 | | (2.1 – 2.3) | 2.3 | (2.2 – 2.3) | 2.3 | | (2.1 – 2.3) | 0.959 |
| Chlorine (mmol/l) | 97 | | (94 - 100) | 97 | (94 - 100) | 97 | | (94 - 100) | 0.696 |
| CPK (IU/l) | 140 | | (76 - 275) | 114 | (73 - 233) | 163 | | (89 - 337) | 0.165 |
| Creatinine (µmol/l) | 104 | | (78 - 135) | 100 | (77 - 214) | 112 | | (78 - 174) | 0.214 |
| Fibrinogen (g/l) | 4.0 | | (3.5 - 4.5) | 4.0 | (3.4 - 4.4) | 4.0 | | (3.5 - 4.6) | 0.805 |
| Haematocrit (%) | 37.9 | | (34.8 - 41.7) | 37.9 | (34.8 - 41.7) | 37.9 | | (35.0 - 41.4) | 0.947 |
| Haemoglobin (g/dl) | 12.7 | | (11.6 - 14.2) | 12.8 | (11.5 - 14.2) | 12.6 | | (11.7 - 14.1) | 0.467 |
| International normalized ratio | 1.0 | | (1.0 - 1.1) | 1.1 | (1.0 - 1.1) | 1.1 | | (1.0 - 1.1) | 0.707 |
| Leucocytes (G/l) | 3.4 | | (2.1 - 5.4) | 5.0 | (3.6 - 6.8) | 5.0 | | (3.4 - 7.9) | 0.869 |
| Lipase (IU/l) | 38.5 | | (24.0 - 58.3) | 39.0 | (25.2 - 57.0) | 34.5 | | (21.7 - 65.6) | 0.162 |
| Lymphocytes (G/l) | 0.6 | | (0.4 - 0.9) | 0.5 | (0.4 - 0.9) | 0.8 | | (0.5 - 1.1) | 0.106 |
| Monocyte to platelet ratio | 3.8 | | (2.5 - 6.7) | 3.8 | (2.5 - 6.7) | 4.0 | | (2.5 - 6.3) | 0.692 |
| Monocytes (G/l) | 0.6 | | (0.4 - 0.8) | 0.6 | (0.4 - 0.8) | 0.6 | | (0.4 - 0.8) | 0.359 |
| Neutrophil to lymphocyte ratio | 5.5 | | (2.8 - 10.7) | 6.5 | (3.2 - 10.8) | 4.2 | | (2.5 - 10.2) | **0.039** |
| Neutrophil to lymphocyte*platelet ratio | 4.3 | | (2.4 - 7.7) | 4.3 | (2.5 - 7.9) | 3.9 | | (1.9 – 9.0) | 0.492 |
| Neutrophil to platelet ratio | 25.8 | | (16.5 - 38.7) | 26.8 | (17.6 - 36.4) | 23.4 | | (15.3 - 45.8) | 0.536 |
| Neutrophils (G/l) | 2.8 | | (1.7 - 4.5) | 3.5 | (2.3 - 5.2) | 3.3 | | (1.9 - 6.1) | 0.731 |
| Phosphate (mmol/l) | 1.0 | | (0.8 - 1.2) | 1.0 | (0.8 – 1.1) | 1.1 | | (0.9 - 1.3) | 0.483 |
| Platelet to lymphocyte ratio | 255 | | (128 - 397) | 277 | (140 - 445) | 222 | | (85 - 365) | 0.169 |
| Platelets (G/l) | 152 | | (100 - 195) | 153 | (105 - 197) | 152 | | (88 - 187) | 0.614 |
| Potassium (mmol/l) | 4.1 | | (3.7 - 4.5) | 4.0 | (3.7 - 4.3) | 4.3 | | (3.8 - 4.7) | **0.008** |
| Prothrombin time (%) | 90 | | (81 - 101) | 89 | (81 - 101) | 91 | | (79 - 102) | 0.513 |
| Sodium (mmol/l) | 136 | | (133 - 138) | 136 | (133 - 138) | 135 | | (132 - 138) | 0.571 |
| Total bilirubin (mg/l) | 7.2 | | (4.9 - 12.3) | 7.9 | (4.9 - 12.1) | 7.1 | | (4.4 - 12.7) | 0.714 |
| Troponin (ng/l) | 0.2 | | (0.1 - 0.6) | 0.2 | (0.1 - 0.3) | 0.2 | | (0.1 - 0.8) | 0.866 |
| Urea (mmol/l) | 6.5 | | (4.8 - 9.6) | 6.3 | (4.8 - 8.4) | 6.9 | | (5.0 - 11.8) | 0.728 |
| Urea to creatinine ratio | 60 | | (49 - 76) | 61 | (51 - 77) | 58 | | (46 - 73) | 0.505 |
| *Biology on hospital stay* (medians, Q_1_-Q_3_) | | |  |  |  |  | |  |  |
| Haemoconcentration | 12 | | (7.7) | 3 | (2.9) | 9 | | (17.0) | **0.003** |
| Platelet nadir (G/l) | 108 | | (42 - 160) | 129 | (51 - 165) | 67 | | (26 - 146) | **< 0.001** |
| Thrombocytopenia (< 100 G/l) | 85 | | (48.3) | 45 | (38.8) | 40 | | (66.7) | **< 0.001** |
| Albumin nadir (g/l), n=64 | 36.7 | | (34.0 - 39.7) | 37.9 | (34.9 - 40.7) | 36.1 | | (32.4 - 38.6) | 0.317 |
| HbA1c (%), n=59 | 7.8 | | (6.6 - 9.0) | 8.1 | (7.1 - 10.7) | 7.5 | | (6.6 - 8.6) | 0.701 |
| Cholesterol total (g/l), n=37 | 3.1 | | (2.6 - 3.5) | 3.1 | (2.6 - 3.7) | 3.1 | | (2.4 - 3.5) | 0.879 |
| Triglycerides (g/l), n=37 | 1.9 | | (1.3 - 2.8) | 1.6 | (1.0 - 2.3) | 2.3 | | (1.4 - 3.1) | 0.243 |
| Peak troponin (ng/l), n=46 | 0.2 | | (0.1 - 0.7) | 0.2 | (0.1 - 0.3) | 0.3 | | (0.1 - 1.1) | 0.231 |
| Peak C Reactive protein (mg/l) | 107 | | (83 - 140) | 101 | (78 - 129) | 122 | | (97 - 199) | **0.011** |
| *Hospital outcomes* |  | |  |  |  |  | |  |  |
| Co-infection | 33 | | (17.9) | 14 | (11.3) | 19 | | (31.7) | **0.001** |
| Hospitalization | 116 | | (63.0) | 63 | (50.8) | 53 | | (88.3) | **< 0.001** |
| LOS (days ; medians, Q_1_-Q_3_) | 6 | | (4 - 9) | 5 | (3 - 7) | 6 | | (4 - 10) | 0.386 |
| Hospitalization in the ICU | 22 | | (12.0) | 3 | (2.4) | 19 | | (31.7) | **< 0.001** |
| Death or need for critical care | 9 | | (5.0) | 0 | (0.0) | 9 | | (15.5) | **< 0.001** |
| *^†^This variable sums the numbers of probable dengue indicators and warning signs. ALT: alanine aminotransferase. AST: aspartate aminotransferase. CPK: creatine phosphor kinase. HbA1c : glycosylated haemoglobin. LOS: length of hospital stay. ICU: intensive care unit. Data are numbers and column percentages, or medians and interquartile ranges (Q1-Q3) when specified. Percentages are compared using chi2 or Fisher’s Exact test, as appropriate. Medians are compared using a non-parametric Brown-Mood k-sample test and when it was significant, further with median and quartile regressions. * p< 0.05, ** p<0.01 and *** p<0.001 for the less significant of both quantile regressions (no * means that at least one quantile regression is not significant).* | | | | | | | | | |
